# Noninvasive brain stimulation combined with evidence-based psychotherapy for psychiatric disorders: A meta-analysis of optimal implementation parameters

**DOI:** 10.64898/2026.02.19.26346650

**Authors:** Lysianne Beynel, Eva Wiener, Neil Baker, Ethan Greenstein, Andrada D. Neacsiu, Eudora Jones, Brian Gindoff, Sunday M. Francis, Cecilia Neige, Marine Mondino, Simon W. Davis, Bruce Luber, Sarah H. Lisanby, Zhi-De Deng

## Abstract

Evidence-based psychotherapies are first-line treatments for psychiatric disorders, yet response rates remain suboptimal. Noninvasive brain stimulation (NIBS) may augment psychotherapy by modulating treatment-engaged circuits. We conducted a systematic review and meta-analysis of randomized controlled trials comparing active NIBS plus evidence-based psychotherapy versus sham NIBS plus psychotherapy. Following Cochrane methods, we searched six databases through February 2025, screening 1,017 records. Twenty-eight trials (31 treatment arms; 1,506 participants) met inclusion criteria. Active NIBS combined with psychotherapy produced significantly greater symptom improvement than sham NIBS with psychotherapy (standardized mean difference = -0.38, 95% confidence interval [-0.68, -0.08]), with substantial heterogeneity. Moderator analyses revealed critical implementation parameters: repetitive transcranial magnetic stimulation (rTMS) showed significant benefit while transcranial direct current stimulation did not. Non-concurrent delivery—stimulation before or after psychotherapy sessions—was significantly effective, whereas concurrent administration was not. Among psychotherapy modalities, cognitive behavioral therapy combined with NIBS produced significant benefit. Human-delivered psychotherapy, but not computerized formats, significantly enhanced outcomes. By diagnosis, significant effects were observed only for anxiety disorders. Secondary analyses revealed significant anxiety symptom reduction specific to rTMS. Treatment integrity was under-reported: only 39.3% of studies used fully manualized protocols and 10.7% documented therapist adherence. Non-concurrent rTMS paired with human-delivered, manualized cognitive behavioral therapy emerges as the most effective strategy, particularly for anxiety disorders. These findings provide an evidence-based framework for optimizing combined treatment protocols and highlight the need for standardized psychotherapy fidelity monitoring in future trials.

## 1 Introduction

Psychiatric disorders impose a large burden of disability and economic loss worldwide (Arias et al., 2022). Evidence-based psychotherapies (EBPs), typically delivered in manualized formats and supported by empirical evidence, are first-line treatments across diagnoses (APA Presidential Task Force on Evidence-Based Practice, 2006; Cook et al., 2017). In posttraumatic stress disorder (PTSD), clinical guidelines from the U.S. Department of Veterans Affairs and Department of Defense strongly recommend manualized EBPs such as prolonged exposure, cognitive processing therapy, and eye movement desensitization and reprocessing over medications when feasible (Schnurr et al., 2024). In obsessive–compulsive disorder (OCD) and body dysmorphic disorder, international guidelines designate exposure and response prevention as the therapy of first choice (National Institute for Health and Care Excellence, 2005). In adult major depressive disorder (MDD), psychotherapy families, including cognitive behavioral therapy (CBT), behavioral activation, interpersonal psychotherapy, and problem-solving therapy, are efficacious and broadly comparable, with benefits that can persist up to 12 months (Cuijpers et al., 2021). Despite this efficacy, short-term response to psychotherapy in MDD is limited to approximately 40% and many do not remit (Cuijpers et al., 2021). Effects vary substantially across trials, and durability is limited: across randomized trials with long-term follow-up, relapse after psychotherapy occurs in roughly 39% of patients (Steinert et al., 2014). Taken together, these data establish EBPs as foundational yet underscore the need for strategies that are more targeted, more durable, and more adaptable to individual profiles, including potentially synergistic combination and/or sequencing with noninvasive brain stimulation (NIBS).

NIBS augments EBPs by directly modulating treatment-engaged circuits with greater focality and typically fewer systemic adverse effects compared with pharmacotherapies (Regenold et al., 2022). Two NIBS modalities are in clinical use: transcranial magnetic stimulation (TMS, including repetitive TMS [rTMS] and theta burst stimulation [TBS]) and transcranial electrical stimulation (tES). TMS generates intracranial electric fields via rapidly changing magnetic fields, which can depolarize neurons and modulate cortical network excitability and plasticity. In the U.S., TMS is cleared for MDD, OCD, smoking cessation, and migraine. For OCD specifically, the FDA-cleared protocol is adjunctive and incorporates brief, individualized symptom provocation immediately before each session (Carmi et al., 2019). tES, which encompasses transcranial direct current stimulation (tDCS), transcranial alternating current stimulation, and transcranial random noise stimulation, perturbs membrane potentials with weak currents and alters cortical excitability (Reed and Cohen Kadosh, 2018). Meta-analytic evidence supports tES superiority over sham for several psychiatric disorders, including MDD, OCD, PTSD, and anxiety disorders (Kalu et al., 2012; Xie et al., 2024). In December 2025, the U.S. FDA approved a home-delivered tDCS device for the treatment of moderate to severe major depressive disorder as monotherapy or adjunctive therapy (Bikson et al., 2026). Because psychotherapies harness learning-dependent plasticity (Craske et al., 2014; Hofmann et al., 2012), temporally aligning stimulation with therapeutic learning episodes provides a biologically grounded rationale for combining these modalities to amplify clinical gains.

The integration of NIBS with psychotherapy is a multimodal approach designed to exploit neural plasticity and the principle of state-dependency to optimize clinical outcomes (He et al., 2022; Saccenti et al., 2024; Sathappan et al., 2019). By time-locking brain stimulation with in-session psychotherapy processes, clinicians utilize “functional targeting” or “cortical paired associative stimulation” to synchronize the modulation of disorder-related neural circuits (e.g., prefrontal cortex), while those circuits are simultaneously engaged by cognitive or emotional interventions (Deng et al., 2020; Luber and Lisanby, 2014; Sathappan et al., 2019). This synergy is grounded in the idea that while psychotherapy promotes behavioral and neural plasticity, NIBS directly targets dysfunctional circuitry to facilitate long-term potentiation or depression, potentially producing faster and more durable therapeutic changes (Saccenti et al., 2024).

The current evidence base suggests potential overall benefit of pairing stimulation with psychotherapy, with larger effects dependent upon specific domains and designs. Across disorders, active rTMS paired with psychological interventions significantly improves overall clinical symptoms compared to sham combinations, yielding a small-to-moderate effect size (SMD = 0.31) (Xu et al., 2023). This multimodal approach appears particularly effective for anxiety, demonstrating a large effect size (Cohen’s *d* = -0.82) when brain stimulation is combined with mindfulness-based interventions (Demina et al., 2025). For depressive symptoms, the evidence is more nuanced; while some data show a small-to-medium effect (Cohen’s *d* = -0.24) that fails to reach statistical significance across all populations (Demina et al., 2025), the combination of NIBS and psychosocial therapy shows a significant positive effect specifically for moderate-to-severe depression (SMD = -0.46) (He et al., 2022). The reported magnitude of these benefits varies significantly based on the trial design and control conditions used. Uncontrolled pretest-posttest comparisons of active combinations show a very large therapeutic effect (Hedges’ *g* = -1.91), whereas controlled comparisons against sham rTMS paired with active non-pharmacological methods result in a medium effect size (Hedges’ *g* = -0.55) (Giron et al., 2025). Furthermore, when compared directly against NIBS monotherapy, the combined approach demonstrates a large and significant effect in reducing depression severity (SMD = -0.84) (He et al., 2022). Beyond mood regulation, integrating rTMS with cognitive training produces significant enhancements in global cognition (SMD = 0.45) (Xu et al., 2023). The augmentation effect of rTMS on CBT also becomes statistically significant when treatments include ten or more sessions (SMD = 0.21) (Xu et al., 2023). While most findings are small-to-moderate, some pilot research has reported exceptional outcomes, such as tDCS paired with positive psychotherapy reaching a very large effect size (Cohen’s *d* = 1.99) compared to stimulation alone (Kochanowski et al., 2024). However, active combinations have not yet consistently outperformed active rTMS when compared against protocols using sham psychological interventions (Giron et al., 2025; He et al., 2022).

Despite the therapeutic promise, the integration of NIBS and psychotherapy faces substantial feasibility and technical hurdles. Concurrent administration can be difficult; for example, the percussive noise and somatic sensations of TMS can hinder mindfulness practice, leading to increased anxiety and high dropout rates (Cavallero et al., 2021). Attrition is a significant concern, with some trials reporting up to a 40% loss to follow-up (Demina et al., 2025; Xu et al., 2023). Timing remains a primary uncertainty; optimal temporal alignment between stimulation and psychotherapy is not established, and protocols currently use three strategies: priming (NIBS before psychotherapy to increase plasticity), online/synergistic (NIBS during psychotherapy to exploit state dependence), and consolidation (NIBS after psychotherapy to stabilize learning). Another challenge concerns psychotherapy treatment integrity, which consists of therapist adherence to the prescribed theoretical model and the competence to deliver those techniques skillfully and appropriately (Esposito et al., 2024). Systematic reviews indicate that treatment integrity is an important dimension associated with favorable clinical outcomes (Esposito et al., 2024; Power et al., 2022); however, this factor is not always systematically accounted for or reported in combination studies or meta-analyses. For example, in some combined protocols, the psychological component is reduced to audio-guided or modified versions that lack the substantive clinician–patient relationship necessary to navigate the negative emotional states or psychological discomfort that may arise during stimulation (Cavallero et al., 2021). The efficacy of combined treatments may be further blunted by external factors, such as the concurrent use of benzodiazepines or antipsychotics, which can interfere with brain response to stimulation (Kochanowski et al., 2024). Finally, widespread implementation remains limited by the high cost and time commitment required for multi-session combined protocols. Taken together, these issues argue for larger, standardized randomized controlled trials to determine feasible schedules and optimal timing, and ensure psychotherapy integrity (Saccenti et al., 2024; Tatti et al., 2022; Xu et al., 2023).

The present meta-analysis quantifies the efficacy of NIBS delivered adjunctively with EBPs in psychiatric populations and tests moderators with clinical and mechanistic salience. We restrict inclusion to clinical psychiatric samples and EBPs. We model moderators spanning NIBS factors (modality: rTMS, tES; timing of stimulation relative to therapy) and psychotherapy factors (treatment modality; treatment integrity), as well as diagnostic category and treatment-resistance status. Our central analysis evaluates whether the temporal relationship between stimulation and psychotherapy modulates effect size. By isolating these parameters, we seek to generate practical recommendations for protocol design and a mechanistic account of how NIBS augments psychotherapy to support durable remission. Our goal is an actionable framework for when and how to combine NIBS with EBPs to improve durability and remission in psychiatric care.

## 2 Methods

This quantitative meta-analysis was performed according to the recommendations of the Cochrane group. Procedures included a comprehensive database search, duplicate removal, dual independent screening at title/abstract and full-text stages according to prespecified inclusion/exclusion criteria, quality and risk-of-bias assessments, data extraction, and quantitative synthesis. The protocol was registered with the International Prospective Register of Systematic Reviews (PROSPERO; registration number CRD42024570287).

### 2.1 Literature search

On February 5, 2025, one investigator (EW) executed searches in the following databases: Scopus, Web of Science, PubMed, Embase, the Cochrane database, and PsycINFO, without date limits and restricted to English-language records. Our chosen keywords for the database search were: (“psychotherapy” OR “psychological intervention”) AND (“noninvasive brain stimulation” OR “NIBS” OR “repetitive transcranial magnetic stimulation” OR “rTMS” OR “transcranial electric stimulation” OR “TES” OR “transcutaneous auricular vagus nerve stimulation” OR “taVNS” OR “transcranial direct current stimulation” OR “tDCS” OR “transcranial focused ultrasound” OR “tFUS” OR “non-invasive brain stimulation” OR “TMS” OR “transcranial magnetic stimulation” OR “TBS” OR “theta burst stimulation” OR “transcranial biophotomodulation” OR “tPBM” OR “transcranial alternating current stimulation” OR “tACS” OR “transcranial random noise stimulation” OR “tRNS” OR “temporal interference stimulation” OR “TI”) AND (“randomized controlled trial” OR “RCT”).

### 2.2 Eligibility criteria

The following selection criteria were applied to papers identified by our search, and based on the Population, Intervention, Comparison, Outcome and Settings framework (PICOS) (Moher et al., 2009). The following criteria were used:

#### 2.2.1 Participants

Males and females aged 15 years or older with psychiatric disorders (including mood disorders, anxiety disorders, psychotic disorders, trauma and stressor related disorders, substance use disorders, eating disorders, impulsive/compulsive disorders, dissociative disorders, somatoform disorders, conduct disorders, and personality disorders); or neurodevelopmental disorders (including autism spectrum disorder and attention-deficit/hyperactivity disorder). Studies in which patients were not formally diagnosed with a DSM-5 disorder were excluded. Ultimately, no studies with neurodevelopmental disorders as the diagnostic category of interest met our inclusion criteria.

#### 2.2.2 Intervention

The included interventions involved NIBS combined with evidence-based psychotherapy. Evidence-based psychotherapy refers to scientifically validated treatments tailored to clients’ individual needs. The National Institute of Mental Health emphasizes that rigorous research has shown that evidence-based therapies reduce psychopathology severity and improve quality of life effectively, and these therapies are generally supported by at least two randomized controlled trials. Studies including cognitive training in tandem with NIBS were not considered to include evidence-based psychotherapy and were excluded.

#### 2.2.3 Comparison

Studies in which the experimental condition was active NIBS with active evidence-based psychotherapy, and the control condition sham NIBS with active evidence-based psychotherapy were included. Studies in which the control condition was sham NIBS alone without psychotherapy, or active NIBS without psychotherapy, or in which no stimulation was considered sham NIBS were excluded.

#### 2.2.4 Outcome

Psychopathology symptoms reported in original studies as the primary or secondary outcome were included. If means and standard deviations were not reported and could not be accessed when contacting the authors, the study was excluded. Studies that did not include psychopathology improvement as an outcome measure (i.e., only included changes in cognitive function or physiological characteristics) were excluded.

#### 2.2.5 Study design

We included randomized controlled trials (including parallel group and crossover designs). Reviews, case report studies, and re-analyses of the same dataset were excluded.

### 2.3 Title, abstract, and full text screening

After deduplication (EW), two independent screeners (EW, NB) evaluated titles/abstracts against eligibility criteria. Discrepancies were discussed and resolved with the rest of the team, and inter-rater reliability was assessed across the corpus. Next, the included studies underwent a full-text screening, performed independently by the same two investigators. Discrepancies were also discussed and resolved with the rest of the team, and inter-rater reliability was assessed.

### 2.4 Clinician screening to evaluate fidelity to evidence-based psychotherapy

Three clinicians (AN, EJ, BG) independently screened the included articles based on the following criteria: 1) the presence of a valid psychiatric diagnosis in participants, as defined by the DSM-5 and confirmed through the use of appropriate evidence-based rating scales; 2) whether the psychotherapy was evidence-based, meaning it followed a treatment manual and had at least two randomized controlled trials supporting its efficacy (Chambless and Hollon, 1998; Tolin et al., 2015); and 3) the way in which psychotherapy was administered, which could be computerized, administered by a human, or a combination of both. In the absence of evidence-based treatments, the clinicians rated whether some evidence-based elements from an evidence-based treatment were included in the type of psychotherapy provided. If no elements of evidence-based treatments were administered, the study was excluded. Additionally, the clinicians recorded the time spent on therapeutic tasks and evaluated whether adherence to the therapy protocol was assessed—that is, whether the study explicitly stated that adherence measurement was conducted. After the screening, all clinicians and the research team convened to discuss and resolve discrepancies.

### 2.5 Quality assessment

The Cochrane risk-of-bias tool 2 (RoB 2) was used to assess the possibility of bias presented by the primary outcome in each of the 28 randomized controlled trials examined (Sterne et al., 2019). The RoB 2 is an adapted version of the original Cochrane risk-of-bias tool that addresses user- and expert-identified limitations, advances its methodology, and includes structured signaling questions and an algorithm to provide evaluators with suggested risk levels (Higgins et al., 2011; Higgins et al., 2019; Sterne et al., 2019). RoB 2 signaling questions span five domains: bias due to the randomization process, deviations from intended interventions, missing outcome data, measurement of the outcome, and selection of the reported result. Each signaling question can be answered in one of five ways: “Yes,” “Probably yes,” “Probably no,” “No,” or “No information.” Each domain’s respective signaling questions are ultimately considered together, allowing the evaluator to classify each domain as having a “low” risk of bias, “high” risk of bias, or “some concerns.” Evaluators then decide on the overall risk of bias. Two investigators (LB, EG) thoroughly reviewed the Cochrane guidelines and each signaling question’s criteria (Higgins et al., 2019; Sterne et al., 2019). Evaluators independently completed the RoB 2 for the primary outcome measure of each study, and discrepancies were resolved through a discussion with three additional research team members.

### 2.6 Data extraction

Two investigators (EW, NB) independently extracted relevant data. First, the general characteristics of each study were collected, such as sample size, participant characteristics (age, percentage female, concurrent medications) across active and sham groups, as well as participants’ diagnoses.

Information about NIBS protocols (i.e., NIBS modality, stimulation target and targeting approach), stimulation amplitude (in mA for tES vs. percent of motor threshold for rTMS), stimulation frequency (only for rTMS), stimulation duration, type of sham control, and the NIBS administration schedule (i.e., the number of sessions per week); and psychotherapy (i.e., type of psychotherapy and therapy schedule [number of sessions per week]) were extracted. We also extracted information regarding the timing of NIBS relative to psychotherapy (concurrently or non-concurrently), to draw more complete inferences on the role of brain state in the efficacy of combined psychotherapy-NIBS treatment approaches.

Finally, the same investigators extracted the means and standard deviations of active/sham pre- /post-treatment for: primary outcome, quality of life outcome (if available), executive functioning outcome (if available), anxiety and depression outcome (regardless of diagnosis, if available). If the primary outcome measure was not a psychopathology outcome (e.g., physiological or cognitive), we consulted the corresponding author and selected the most relevant secondary clinical outcome listed.

### 2.7 Categorization of variables

Some variables required categorization to have a large enough sample size per variable for analysis. NIBS modalities used in the studies included tDCS, rTMS, deep TMS (dTMS), and intermittent theta burst stimulation (iTBS). We combined rTMS, dTMS and iTBS into one category called “rTMS.” There were also many distinct psychotherapies utilized across studies, and, via a discussion with the clinicians, three global categories were generated: Cognitive Behavioral Therapy (CBT), mindfulness, and exposure. CBT included any therapies based on cognitive strategies to process thoughts and behaviors. Mindfulness included any therapies described as using mindfulness techniques. Exposure included symptom provocation and exposure and response prevention therapies. Lastly, with the help of the clinicians, we categorized diagnoses included in the studies into the following categories: OCD, depression, anxiety (studies combining the diagnoses of specific fears, acrophobia, social anxiety, and panic disorder), addiction (studies combining nicotine addiction, methamphetamine addiction, and alcohol use disorder), PTSD, transdiagnostic (studies that allowed any DSM-5 disorder), pain (including fibromyalgia, which is listed under somatic symptom disorder in the DSM-5), and personality (including borderline personality disorder [BPD]).

### 2.8 Quantitative Analysis

All statistical analyses were conducted using R (v.4.5.2), with the “meta” and “metafor” packages, which include a collection of functions for conducting meta-analyses (https://CRAN.R-project.org/package=meta). We used the “metacont” function to compute standardized mean differences (SMD) with 95% confidence intervals under a random-effects model. Heterogeneity was summarized with *τ*² and *I*². In cases where standard deviations were unavailable for clinical outcomes (n = 2), we used the pooled standard deviation from all other studies (an average of all available standard deviations). Publication bias was assessed using a funnel plot of the primary outcome measures and Egger’s test (Egger et al., 1997).

#### 2.8.1 Primary analysis

The primary clinical outcome defined by the study was extracted. For studies that had physiological outcomes as their primary measure, the most relevant secondary clinical outcome was used. In the rare event where an increase in clinical outcome score signaled improvement, we inversed the scores to match the direction of change of the rest of the outcomes. Further, subgroup analyses were conducted with the following variables: NIBS modality (tDCS or rTMS), NIBS timing (concurrent or non-concurrent), therapy category (mindfulness, CBT, or exposure), diagnostic category (OCD, depression, anxiety, addiction, PTSD, transdiagnostic, pain, or personality), the degree to which the psychotherapy was evidence-based (i.e., whether it was a fully evidence-based intervention as defined by a manual, or if it only contained partial elements derived from evidence-based practice), and administration method (human, computerized, or mixed). We intended to conduct a subgroup analysis comparing studies where adherence was rated and reported, and studies lacking this information. However, because only one study reported adherence ratings, this analysis could not be conducted. Interactions between variables were also tested: NIBS modality by timing, therapy category by diagnostic category, and therapy category by NIBS modality. Further, if a category, or a combination of categories, did not contain at least three studies, we did not include that category in our subgroup analysis.

#### 2.8.2 Secondary analysis

We performed secondary analyses for the following domains: depression, anxiety (regardless of the diagnosis), executive functioning, and quality of life. Only studies that listed an outcome in each domain were included in each secondary analysis, and, given sample size, we only conducted subgroup analyses within each domain for NIBS modality and NIBS timing.

## 3 Results

### 3.1 Search results

Search criteria yielded 1728 citations. After removing duplicates (n = 711), 1017 articles underwent abstract and title screening, with 31 total disagreements between the two evaluators (3.1%). A total of 668 articles were kept, and then the full text screening was done, with 10 total disagreements between the two evaluators (17.2%). 45 papers passed the full text screening and were sent to the clinicians for rating the level of evidence supporting the psychotherapy, after which 31 papers remained eligible. However, data were missing for three of the papers despite contacting the authors, those publications were not included later in the meta-analysis. Thus, 28 studies were included in the final cohort (**Figure 1**).

**Figure 1.**
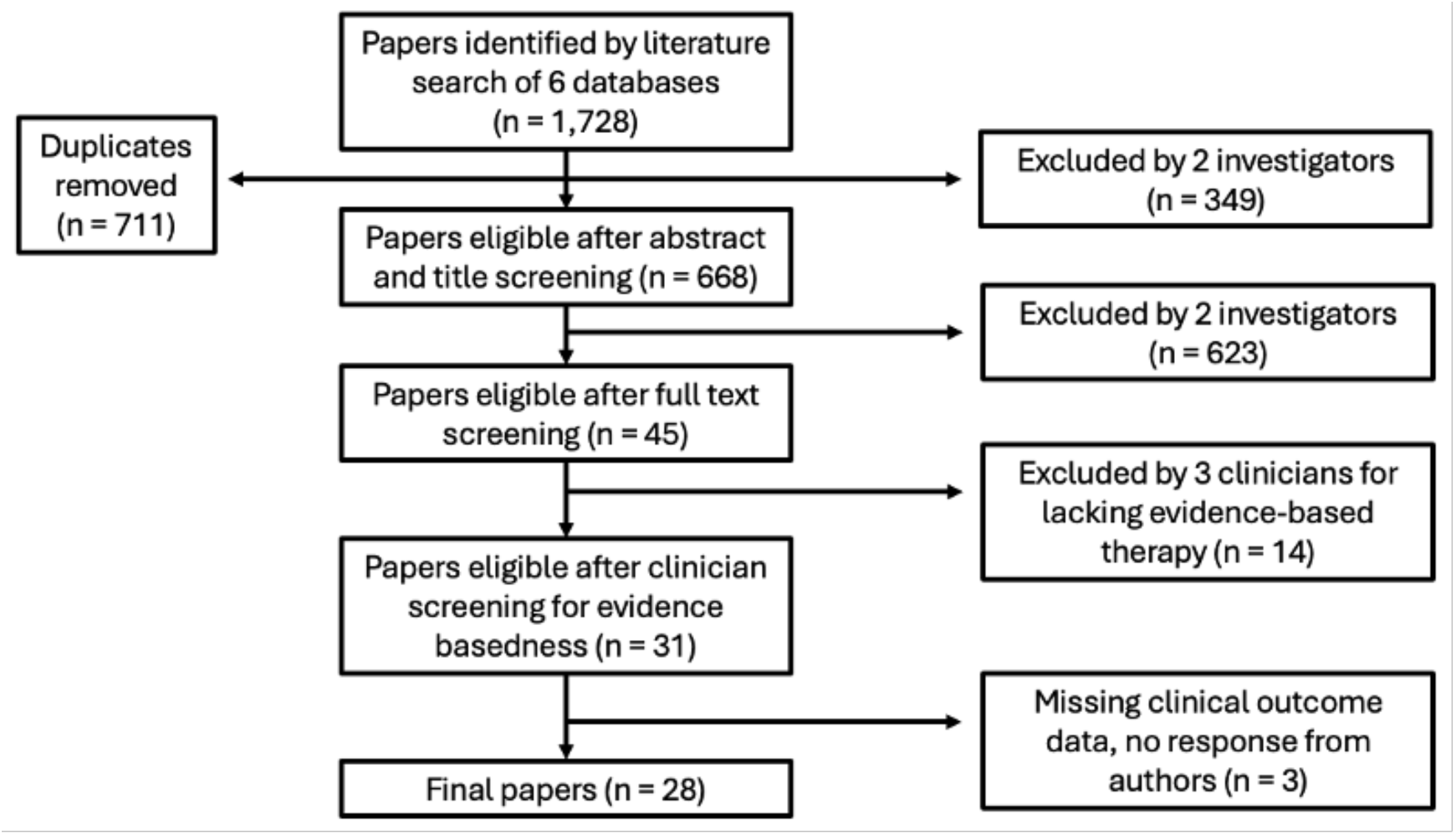
Consort diagram of randomized controlled trials (RCTs) included in the meta-analysis.

**Figure 2.**
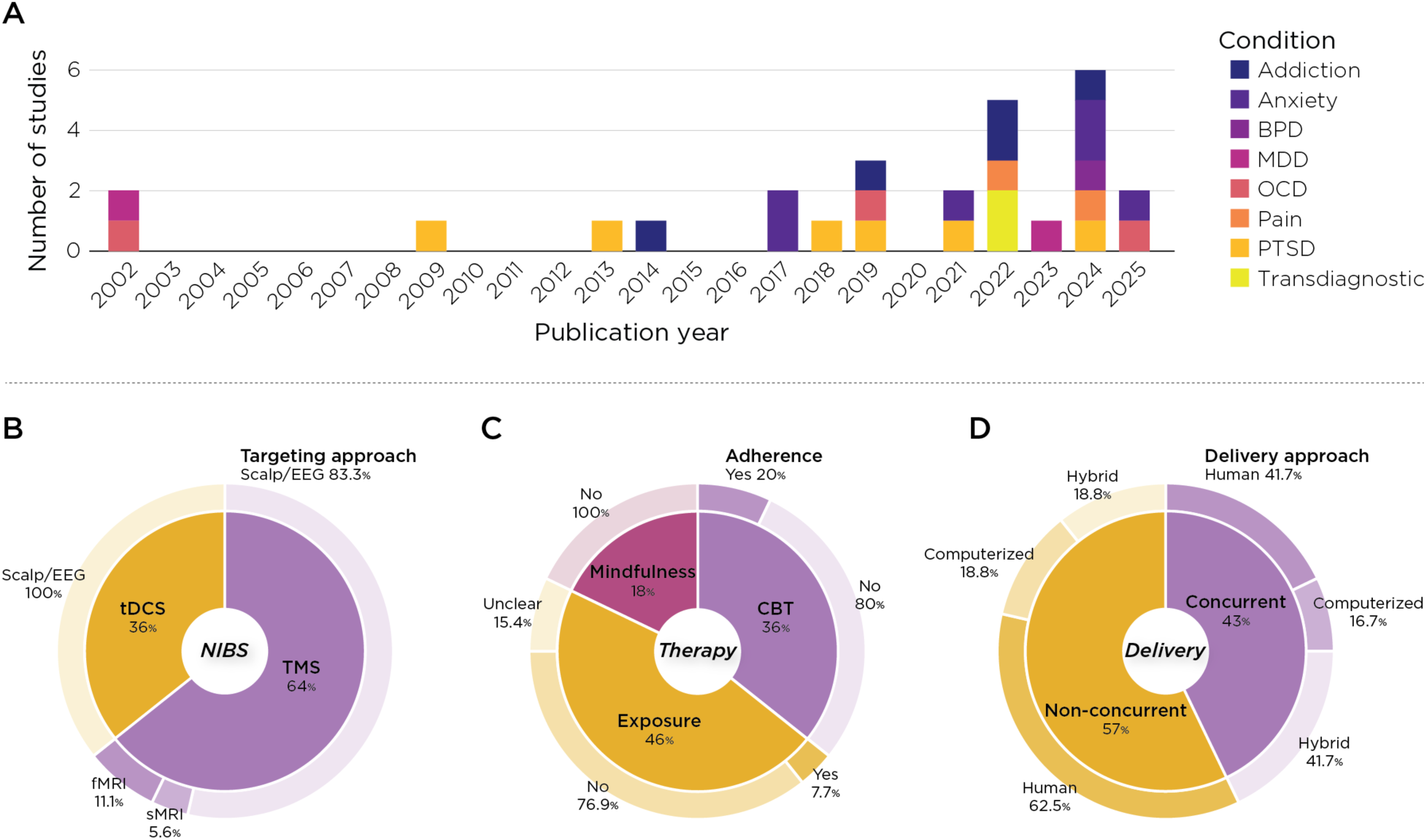
Study landscape and protocol characteristics in trials combining NIBS with psychotherapy. (A) Annual number of included randomized trials by primary diagnostic group (stacked colors: Addiction, Anxiety, BPD, MDD, OCD, Pain, PTSD, Transdiagnostic). (B) NIBS modalities and targeting: TMS accounted for 64% of trials and tDCS for 36%; TMS targeting was predominantly scalp/EEG-based (83.3%), with fewer MRI-guided approaches (fMRI 11.1%, sMRI 5.6%). (C) Psychotherapy categories and integrity reporting: Exposure-based interventions were most common (46%), followed by CBT (36%) and mindfulness (18%). Treatment adherence/competence (“integrity”) was explicitly assessed in only 20% of the CBT trials. (D) Delivery timing and format: Non-concurrent scheduling (stimulation before/after psychotherapy) was more frequent (57%) than concurrent delivery (43%). Psychotherapy was delivered by a human clinician (41.7%), in hybrid formats (41.7%), or computerized (16.7%). Percentages reflect the proportion of studies within the included RCT sample (n=28). Abbreviations: NIBS, noninvasive brain stimulation; TMS, transcranial magnetic stimulation; tDCS, transcranial direct current stimulation; CBT, cognitive behavioral therapy; BPD, borderline personality disorder; MDD, major depressive disorder; OCD, obsessive–compulsive disorder; PTSD, posttraumatic stress disorder.

### 3.2 Descriptive statistics

The following sections provide a general overview of the NIBS parameters, clinical populations and psychotherapies used in combination with NIBS. The following results are expressed as a percentage out of the 28 included studies, except for the stimulation target. As three of the studies (Fitzsimmons et al., 2025; Hu et al., 2022; Neacsiu et al., 2022b) included two active arms, each with different targets, we present the results from 31 total arms across 28 studies. More details about each study are provided in the Supplementary Material.

#### 3.2.1 NIBS

Of the 28 studies identified by our search, only studies using TMS (64.3%) and tDCS (35.7%) have been combined with various psychotherapy approaches in clinical trials matching our criteria. No studies combining tACS, tRNS, or tFUS with psychotherapy were published.

For TMS studies, most used conventional rTMS protocols (72.2%), while some used intermittent theta-burst stimulation (27.8%). To determine the stimulation site, the predominant targeting method was scalp measurement, with or without the 10-20 EEG grid as a reference (83.3%). A minority of studies used fMRI-based targeting (11.1%) or targeting based on an anatomical MRI scan (5.6%). Of the 31 arms in our analysis, rTMS stimulation targets included the left prefrontal cortex (PFC; 38.7%), medial PFC (25.8%), right PFC (22.6%), motor cortex (9.7%), and bilateral PFC (3.2%). As expected, given the FDA-approved label for treatment, the most frequently used stimulation frequency was 10 Hz (44.4%), followed by 5 Hz (22.2%), 1 Hz (11.1%), 20 Hz (11.1%), 18 Hz (5.6%), and 50 Hz (5.6%). The stimulation amplitudes used were 80% rMT (27.8%), 100% rMT (27.8%), 110% rMT (16.7%), and 120% rMT (27.8%).

All studies using tDCS relied on scalp measurements/EEG grid to define their stimulation target. The most common target was either the left PFC (30%) or the medial PFC (30%), followed by the right PFC (20%) and the motor cortex (20%). Most studies used a stimulation amplitude of 2 mA (80%), with only one study using 1.7 mA (10%) and another using 1.5 mA (10%) for increased comfort during stimulation.

#### 3.2.2 Clinical diagnosis, therapy, and combination of therapy with NIBS

Across all studies, the average total sample size was 53.8 ± 33.6 subjects. Clinical populations included patients with anxiety (21.4%), PTSD (21.4%), addiction (17.9%), OCD (10.7%), and pain (10.7%). Only a small percentage of the studies included patients with MDD (7.1%), transdiagnostic emotion dysregulation (7.1%), and BPD (3.6%). Concurrent medication was allowed in most studies (78.6%), with the remaining studies prohibiting medication (10.7%) or not specifying medication status (10.7%). The most common type of therapy used was exposure (46.4%), followed by CBT (35.7%), and mindfulness (17.9%). Across all therapy types, the average total time in therapy was 448.5 minutes (∼7.5 hours), with the average session total being 9.6 sessions. Most studies employed a therapist (53.6%), while some relied on a mix of therapist interaction and computerized elements (28.5%), and two studies used computerized interventions only (7.1%). A minority of studies used a fully evidence-based procedure (39.3%), with most using only elements of an evidence-based protocol (60.7%). This classification was performed via a discussion between clinicians (AN, EJ, BG). Of note, only 3 studies (10.7%) reported adherence measures for the therapeutic protocol used.

Finally, regarding the combination of NIBS with psychotherapy, the majority of studies delivered NIBS right before or right after the psychotherapy intervention (57.1%), with the rest concurrent. Four studies (14.3%) used only one session of NIBS combined with one therapy session, while most of the studies (85.7%) administered multiple sessions of NIBS combined with psychotherapy.

### 3.3 Risk of bias assessment

Two investigators (EG, LB) independently assessed risk of bias using the RoB 2 tool for the primary outcome of each study. Inter-rater agreement was high (80% agreement), with concordant judgments on 112 of 140 ratings. Discrepancies between the two investigators were resolved through group discussion.

Domain-level assessment revealed the following risk profiles: randomization process (Domain 1): 27 low risk (96.4%), 1 some concerns (3.6%); deviations from intended interventions (Domain 2): 21 low risk (75.0%), 7 some concerns (25.0%); missing outcome data (Domain 3): 26 low risk (92.9%), 2 some concerns (7.1%); measurement of outcome (Domain 4): 27 low risk (96.4%), 1 some concerns (3.6%); selection of reported result (Domain 5): 4 low risk (14.3%), 24 some concerns (85.7%). The predominant concern across studies was lack of pre-registration (Domain 5), which affected the majority of trials.

We deviated from the RoB2 algorithm in one specific instance: when lack of pre-registration (Domain 5) was the only concern, we adjusted the overall rating from "some concerns" to "low risk" for 14 studies (50%). This decision reflects that pre-registration is a relatively recent practice and several included trials preceded widespread adoption of this standard. Without this adjustment, the majority of studies would have been classified as having "some concerns" solely based on publication era, obscuring more substantive methodological risks.

Overall, 20 studies (71.4%) were rated as low risk of bias, 6 (21.4%) as some concerns, and 2 (7.1%) as high risk (**Figure 3**).

**Figure 3.**
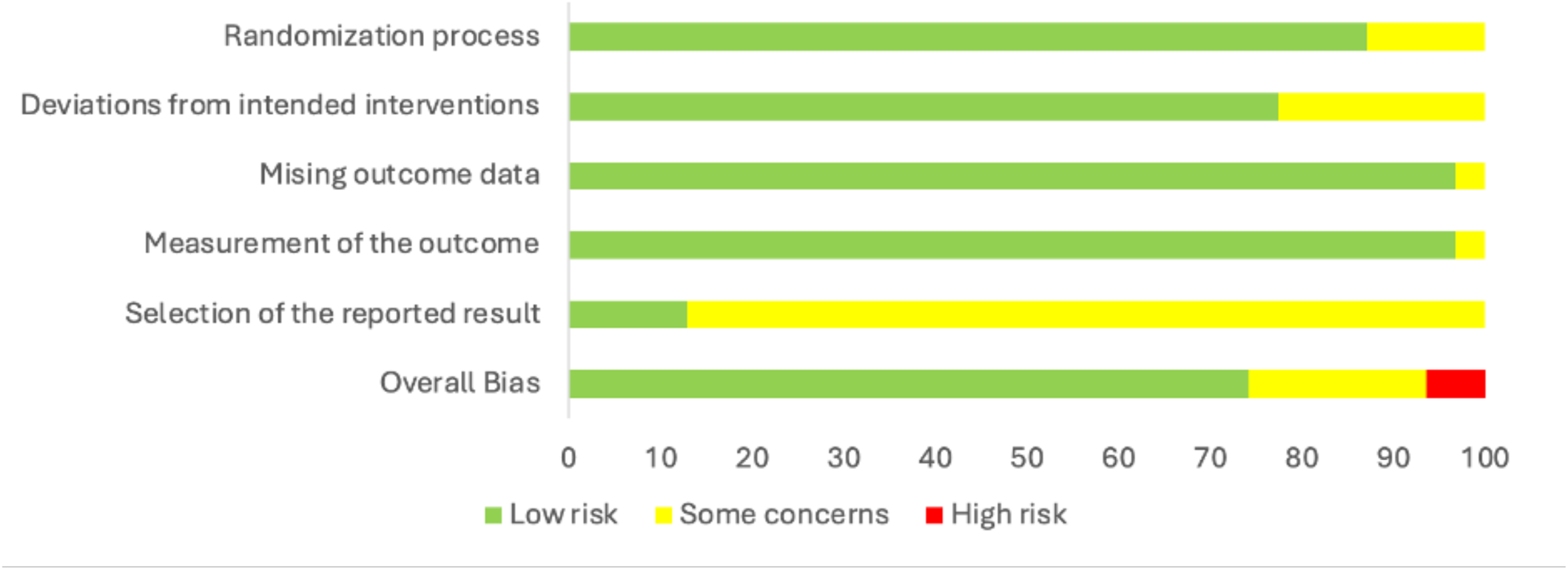
Summary of the risk of bias across the 28 included studies.

### 3.4 Meta-analysis

#### 3.4.1 Primary clinical outcomes

Out of our 28 included papers (and 31 treatment arms, given that three papers included two active arms), 26 reported a clinical outcome as their primary outcome measure, while two reported physiological outcomes as their primary. For those two papers, we consulted the author and selected the most relevant secondary clinical outcome listed. Across all studies, the random-effects meta-analysis significantly greater improvement in primary clinical symptoms after active NIBS with psychotherapy compared to sham NIBS with psychotherapy (SMD = -0.38, 95% CI = [-0.68; -0.08]), with substantial heterogeneity (*I*^2^ = 82.9%). We then conducted subgroup analyses to elucidate differences in clinical outcomes within various categories of interest.

#### 3.4.2 NIBS modality

rTMS had a statistically significant benefit in improving clinical outcomes when paired with psychotherapy (SMD = -0.47, 95% CI = [-0.80; -0.13]), while tDCS did not (SMD = -0.20, 95% CI = [-0.88; 0.47]) (**Figure 4**).

**Figure 4.**
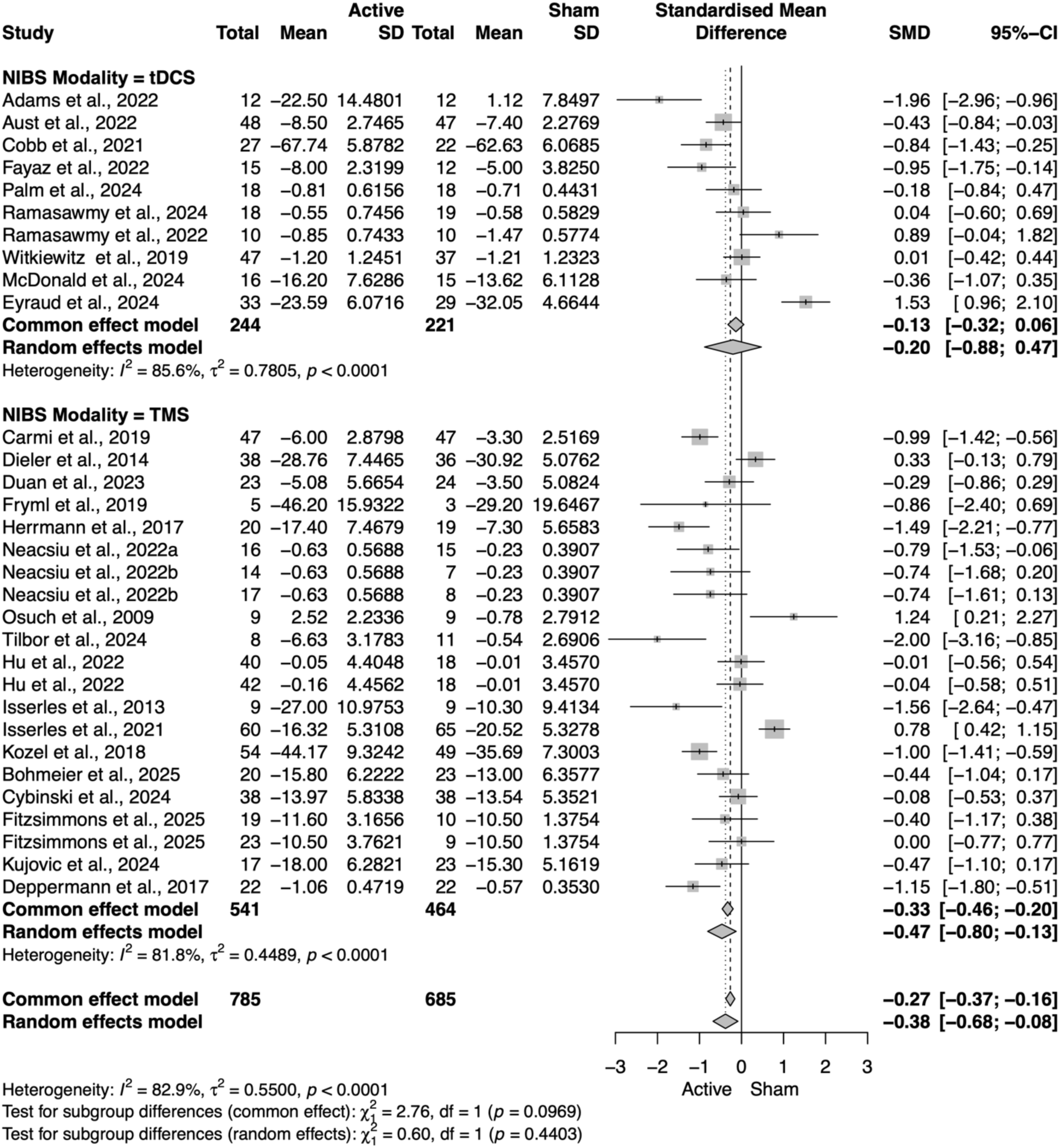
Primary outcome forest plot: Active NIBS+psychotherapy vs. sham NIBS+psychotherapy, stratified by modality. Standardized mean differences (SMD; negative values favor **Active**) are shown for each trial with 95% CIs, pooled within tDCS and rTMS subgroups, and pooled overall using a random-effects model. Both subgroups show a small benefit of combining stimulation with psychotherapy relative to sham+psychotherapy. Heterogeneity is substantial (see *I*² and *τ*²), consistent with variation in protocols, diagnoses, and timing. Column headings report group Ns, means, and SDs used for SMD estimation; studies are weighted by inverse variance. Abbreviations: NIBS, noninvasive brain stimulation; tDCS, transcranial direct current stimulation; rTMS, repetitive transcranial magnetic stimulation; CI, confidence interval.

#### 3.4.3 Diagnostic category

The combination of NIBS and psychotherapy was statistically significant in improving clinical symptoms in those diagnosed with anxiety (SMD = -0.70, 95% CI = [-1.26; -0.14]), but not in those diagnosed with addiction (SMD = -0.07, 95% CI = [-0.48; 0.33]), OCD (SMD = -0.81, 95% CI = [-2.10; 0.49]), pain (SMD = -0.32, 95% CI = [-3.95; 3.32]), or PTSD (SMD = 0.07, 95% CI = [-1.30; 1.45]). The diagnostic categories of depression, personality disorders, and transdiagnostic disorders did not meet the minimum study sample size of three studies and therefore were excluded from this subgroup analysis.

#### 3.4.4 Psychotherapy category

In the subgroup of psychotherapy category (CBT, exposure, and mindfulness), the combination of CBT and NIBS resulted in statistically significant benefit in improving clinical outcomes (SMD = -0.48, 95% CI = [-0.78; -0.17]), while exposure therapy and mindfulness did not (SMD = -0.35, 95% CI = [-0.94; 0.24], SMD = -0.22, 95% CI = [-1.45; 1.01], respectively).

#### 3.4.5 Level of evidence supporting the psychotherapy

Studies that included fully evidence-based psychotherapy had a statistically significant improvement in clinical outcomes for active vs. sham NIBS (SMD = -0.33, 95% CI = [-0.59; - 0.07]), while studies that only used partial elements of evidence-based therapy did not (SMD = - 0.41, 95% CI = [-0.90; 0.08]).

#### 3.4.6 Psychotherapy administration method

Only studies in which psychotherapy is administered exclusively by a human significantly improved clinical outcomes for active vs. sham NIBS (SMD = -0.49, 95% CI = [-0.78; -0.21]). No significant improvement was found with administration solely by a computer (SMD = -0.17, 95% CI = [-1.87; 1.52]) or by a mix of computer and human (SMD = -0.28, 95% CI = [-0.98; 0.43]).

#### 3.4.7 NIBS timing; NIBS timing x NIBS modality

When administered non-concurrently with the psychotherapy (before or after), NIBS had a statistically significant benefit in improving clinical outcomes (SMD = -0.56, 95% CI = [-0.94; - 0.18]), while concurrent NIBS did not (SMD = -0.11, 95% CI = [-0.61; 0.39]). Interestingly, we found that only rTMS given non-concurrently had a statistically significant benefit in improving clinical symptoms (SMD = -0.47, 95% CI = [-0.91; -0.02]). No effects were found with concurrent tDCS (SMD = 0.26, 95% CI = [-0.56; 1.07]), non-concurrent tDCS (SMD = -0.92, 95% CI = [-2.04; 0.20]), or concurrent rTMS (SMD = -0.48, 95% CI = [-1.17; 0.21]) (**Figures 4 and 5**). Unfortunately, the limited number of studies prevented us from testing for an interaction between timing, NIBS modality and each type of psychotherapy, which could have provided stronger insight on this result.

**Figure 5.**
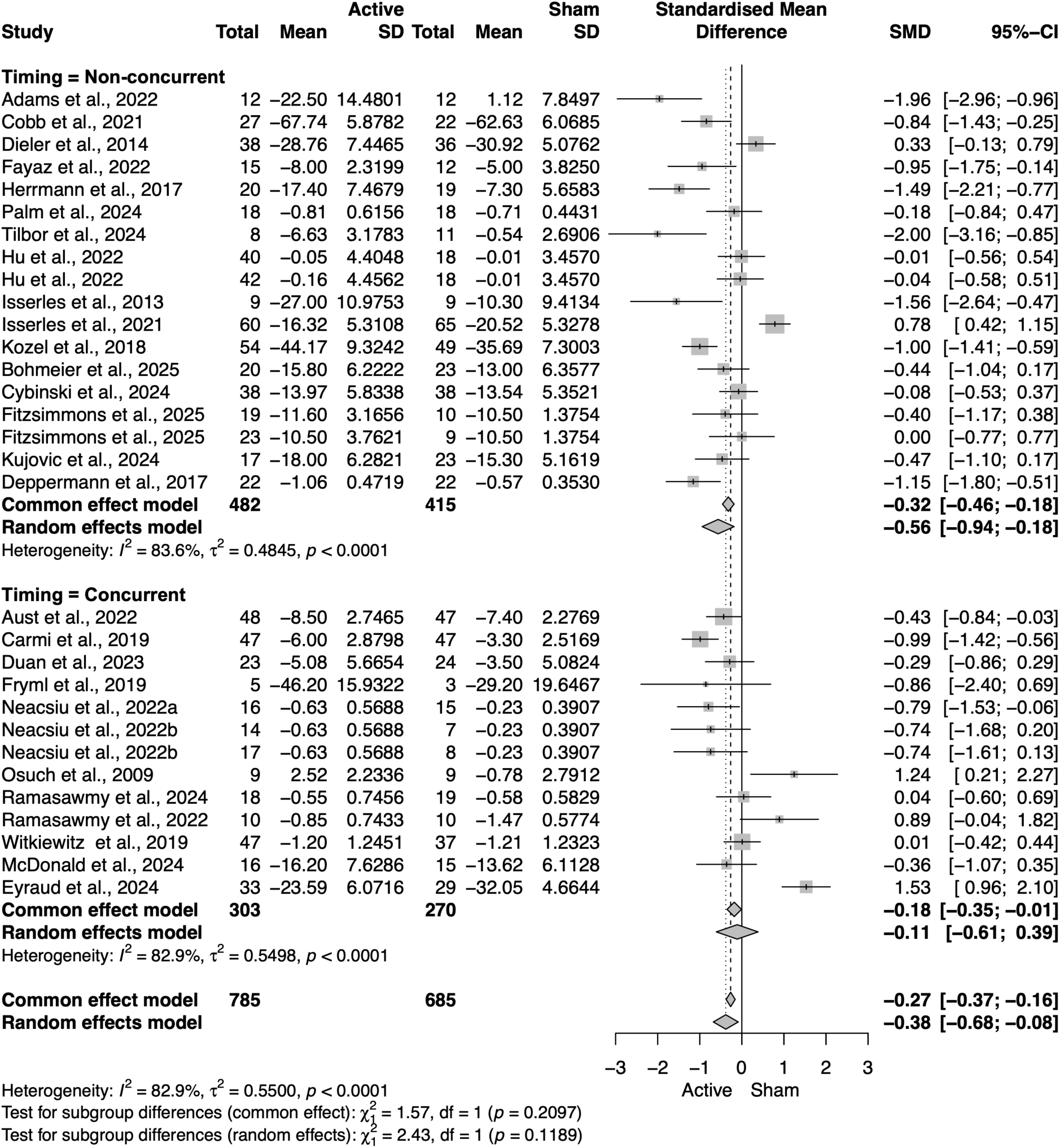
Primary outcome by stimulation–therapy timing. Forest plot for individual trials, pooled within Non-concurrent (stimulation delivered before/after psychotherapy) and Concurrent (stimulation during psychotherapy) subgroups and overall (random-effects). The Non-concurrent subgroup shows a significant small-to-moderate symptom reduction relative to sham+psychotherapy, whereas the Concurrent subgroup’s pooled effect is not statistically significant.

**Figure 6.**
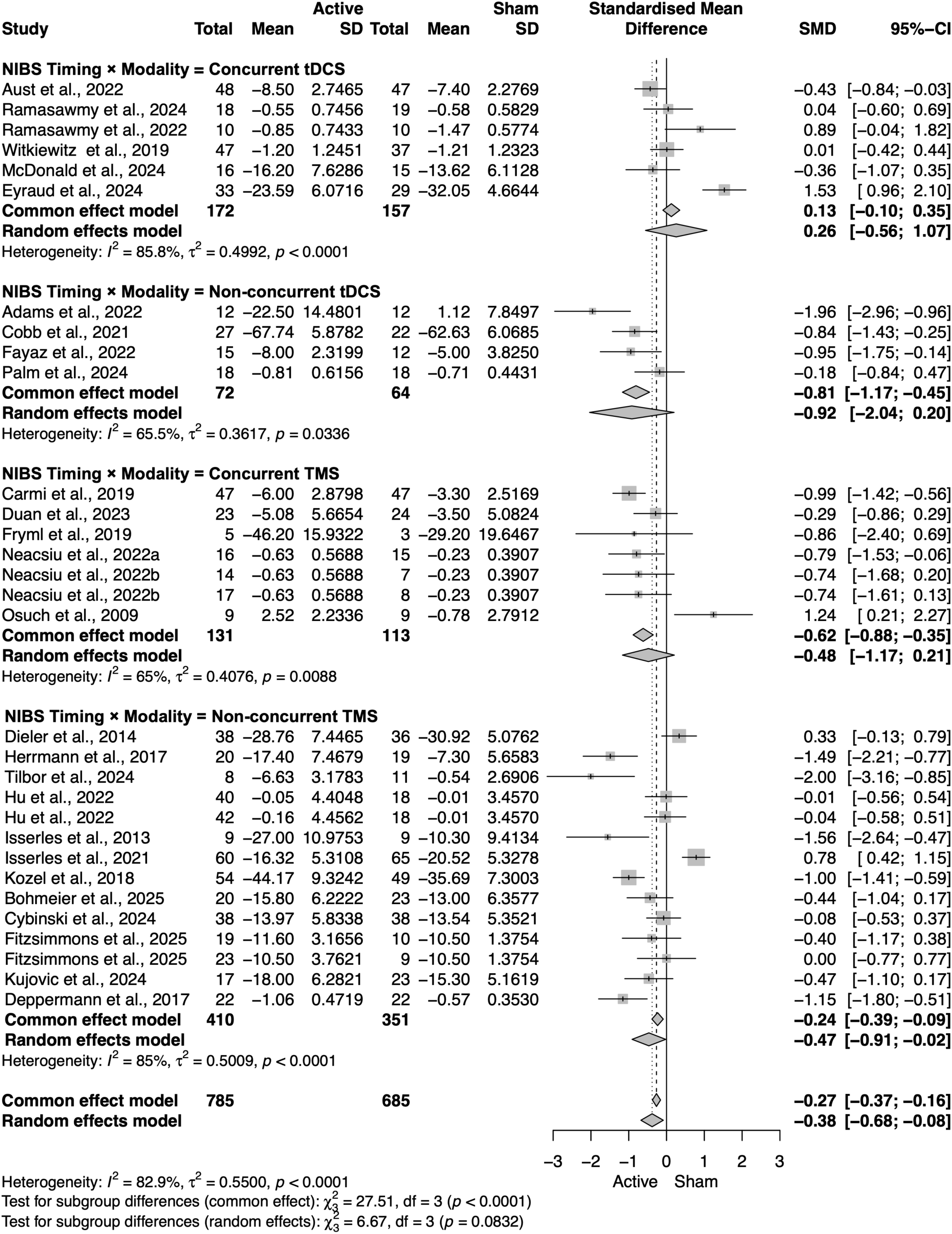
Primary outcome by timing x modality. Forest plot of SMD for individual trials, pooled within four subgroups: Concurrent tDCS, Non-concurrent tDCS, Concurrent rTMS, and Non-concurrent rTMS. Under random effects, only Non-concurrent rTMS shows a significant pooled benefit; the other subgroups are not significant.

#### 3.4.8 NIBS modality × psychotherapy category

We additionally explored the combination of NIBS modality with psychotherapy category. When CBT was administered with rTMS, there was a statistically significant benefit in improving clinical symptoms (SMD = -0.48, 95% CI = [-0.88; -0.07]), while CBT with tDCS (SMD = -0.47, 95% CI = [-1.31; 0.36]), exposure therapy with tDCS (SMD = -0.38, 95% CI = [-2.69; 1.93]), exposure therapy with rTMS (SMD = -0.35, 95% CI = [-0.99; 0.29]), and mindfulness with tDCS (SMD = 0.22, 95% CI = [-11.91; 9.79]) did not. There were only two studies that utilized the combination of mindfulness with rTMS, and those were excluded from this subgroup analysis.

### 3.5 Summary of all results for the primary outcome measures

Given the number of comparisons, we provide in **Table 1** a summary of all the results from the primary outcomes.

**Table 1.** Standardized mean differences (SMD, active vs. sham) for various NIBS modalities, clinical diagnoses, and combined therapeutic interventions. SMD represent the central effect size, flanked by 95% Confidence Intervals (CI). Values in bold denote statistical significance (where the CI does not cross zero), indicating a superior outcome for active stimulation over sham.

| Category | Label | SMD | Lower CI | Upper CI | Significance |
| --- | --- | --- | --- | --- | --- |
| NIBS type | tDCS | -0.20 | -0.88 | 0.47 | Not Significant |
|  | rTMS | -0.47 | -0.80 | -0.13 | **Significant** |
| Diagnosis | Addiction | -0.08 | -0.48 | 0.33 | Not Significant |
|  | Anxiety | -0.70 | -1.26 | -0.14 | **Significant** |
|  | Depression | -0.39 | -1.24 | 0.47 | Not Significant |
|  | OCD | -0.81 | -2.10 | 0.49 | Not Significant |
|  | Pain | -0.32 | -3.95 | 3.32 | Not Significant |
|  | Personality | -0.47 | -1.10 | 0.17 | Not Significant |
|  | PTSD | 0.08 | -1.30 | 1.45 | Not Significant |
|  | Transdiagnostic | -0.76 | -0.84 | -0.68 | **Significant<sup>†</sup>** |
| Therapy | CBT | -0.48 | -0.78 | -0.17 | **Significant** |
|  | Exposure | -0.35 | -0.94 | 0.24 | Not Significant |
|  | Mindfulness | -0.22 | -1.45 | 1.01 | Not Significant |
| Evidence-based | No | -0.41 | -0.90 | 0.08 | Not Significant |
|  | Yes | -0.33 | -0.59 | -0.07 | **Significant** |
| Therapy delivery | Computerized | -0.18 | -1.87 | 1.52 | Not Significant |
|  | Mixed | -0.28 | -0.98 | 0.43 | Not Significant |
|  | Human | -0.50 | -0.78 | -0.21 | **Significant** |
| Timing | Not concurrent | -0.56 | -0.94 | -0.18 | **Significant** |
|  | Concurrent | -0.11 | -0.61 | 0.39 | Not Significant |
| Timing × NIBS | Concurrent tDCS | 0.26 | -0.56 | 1.07 | Not Significant |
|  | Non-concurrent tDCS | -0.92 | -2.04 | 0.20 | Not Significant |
|  | Concurrent rTMS | -0.48 | -1.17 | 0.21 | Not Significant |
|  | Non-concurrent rTMS | -0.47 | -0.91 | -0.02 | **Significant** |
| Therapy × NIBS | CBT with tDCS | -0.48 | -1.31 | 0.36 | Not Significant |
|  | CBT with rTMS | -0.48 | -0.88 | -0.07 | **Significant** |
|  | Exposure with tDCS | -0.38 | -2.69 | 1.93 | Not Significant |
|  | Exposure with rTMS | -0.35 | -0.99 | 0.29 | Not Significant |
|  | Mindfulness with tDCS | 0.22 | -0.88 | 1.31 | Not Significant |
|  | Mindfulness with rTMS | -1.06 | -11.91 | 9.79 | Not Significant |
| Therapy × Diagnosis | CBT for Addiction | -0.11 | -0.65 | 0.44 | Not Significant |
|  | CBT for Transdiagnostic | -0.76 | -0.84 | -0.68 | **Significant<sup>†</sup>** |
|  | Exposure for Anxiety | -0.61 | -1.28 | 0.06 | Not Significant |
|  | Exposure for OCD | -0.81 | -2.10 | 0.49 | Not Significant |
|  | Exposure for PTSD | 0.32 | -1.35 | 1.98 | Not Significant |
|  | Mindfulness for Pain | -0.22 | -2.09 | 1.66 | Not Significant |

<sup>†</sup> Significant but aggregated across only two studies

### 3.6 Publication bias and heterogeneity

Visual inspection of the funnel plot did not suggest asymmetry, and Egger’s test was non-significant (p > 0.05), indicating no evidence for small-study effects (**Figure 8**).

**Figure 7.**
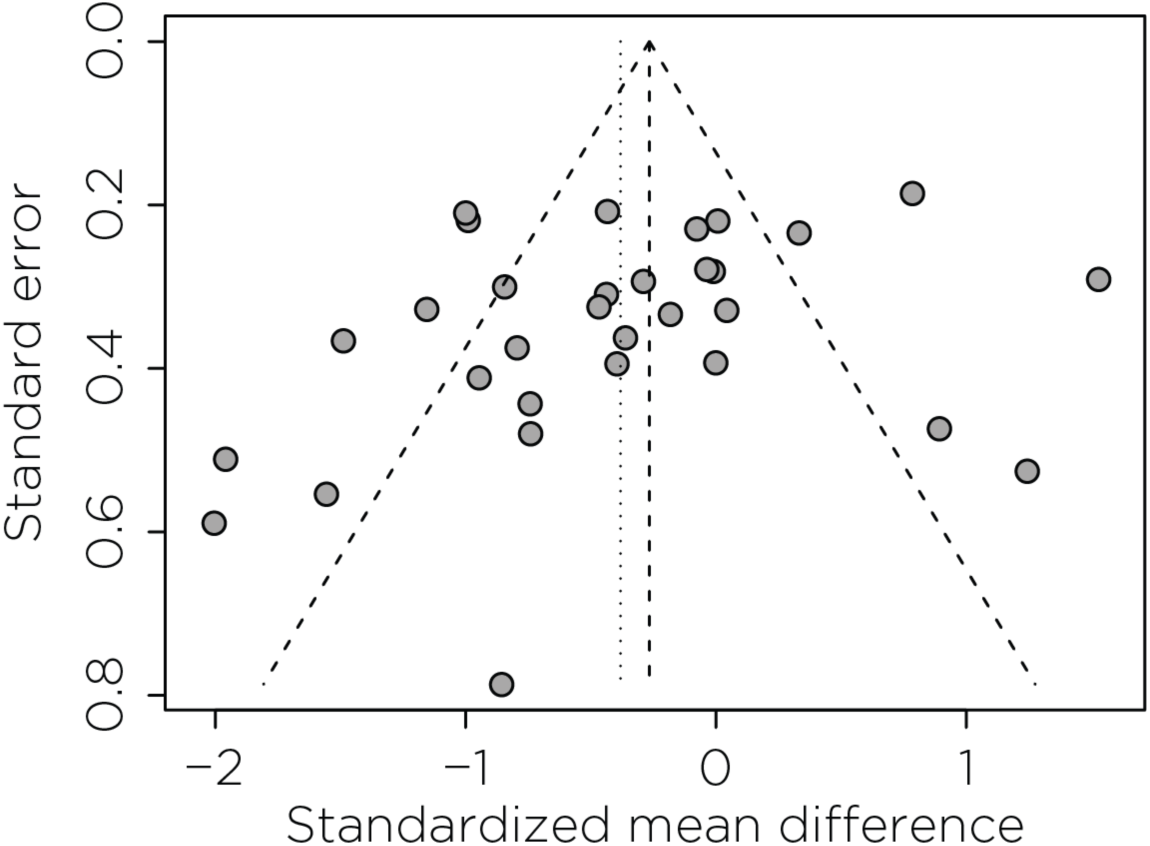
Funnel plot for small-study effects in the primary analysis. Scatter of individual studies’ SMD against their standard errors; negative SMD indicates symptom improvement favoring Active NIBS+psychotherapy. The vertical dashed line marks the pooled random-effects estimate; dotted lines denote the pseudo 95% confidence limits forming the funnel.

**Figure 8.**
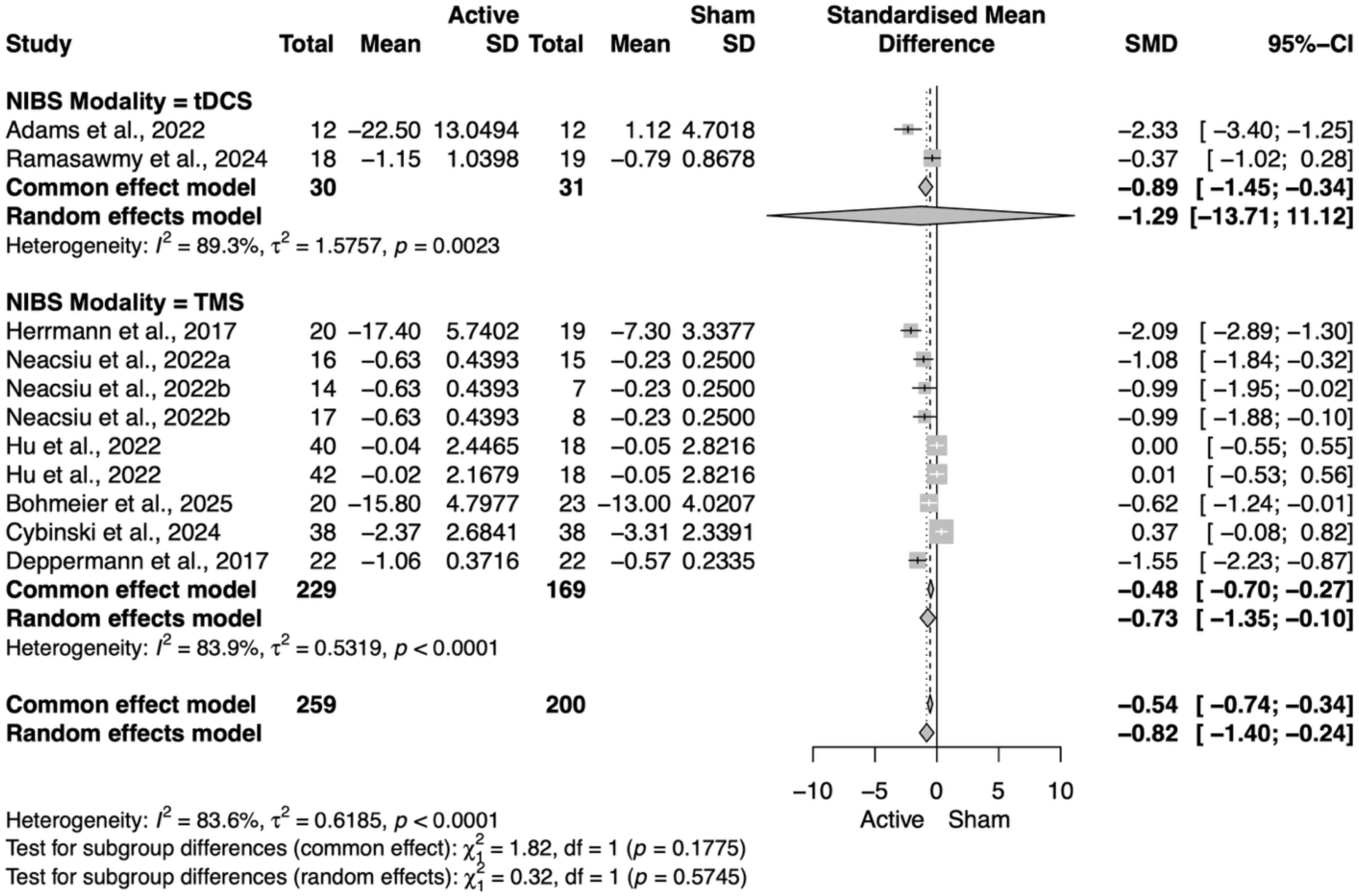
Forest plot for subcategory of NIBS modality on anxiety outcomes.

### 3.7 Executive functioning and quality of life outcomes

No significant results were found for active compared to sham NIBS with psychotherapy in improving executive functioning (n = 9 studies, SMD = 0.12, 95% CI = [-0.45; 0.70]) or quality of life (n = 9 studies, SMD = 0.16, 95% CI = [-0.11; 0.42]).

### 3.8 Other outcomes

We sought to investigate changes in fMRI BOLD activity, as neuroimaging outcomes could provide mechanistic insights into how combined treatment alters neural circuits. However, only four studies collected fMRI at any timepoint, precluding meta-analytic synthesis. Two used fMRI for the purpose of individualized NIBS targeting (Cybinski et al., 2024; Neacsiu et al., 2022a), while the other two explored changes in network activation post-NIBS and psychotherapy (Adams et al., 2022; Fitzsimmons et al., 2025). Given the lack of studies and the variability in the tasks employed, we were unable to conduct this analysis.

Finally, we conducted an analysis of studies that included depression and anxiety outcome measures, independently of the diagnosis. We did not find any significant results for active compared to sham NIBS with psychotherapy in improving depression symptoms (SMD = -0.08, 95% CI = [-0.35; 0.19]). However, we did find an overall statistically significant improvement in anxiety scales for active NIBS with psychotherapy compared to sham NIBS with psychotherapy (SMD = -0.82, 95% CI = [-1.40; -0.24]). With subgroup analysis, we found that this improvement in anxiety measures was specific to rTMS (SMD = -0.73, 95% CI = [-1.35; -0.10]) (**Figure 9**). We additionally explored the subgroup of NIBS timing and found that, when administered concurrently, the combination of NIBS and psychotherapy was statistically significant in improving anxiety symptoms (SMD = -0.80, 95% CI = [-1.38; -0.23]), while non-concurrent NIBS with psychotherapy was not significant (SMD = -0.83, 95% CI = [-1.83; 0.17]) (**Figure 10**).

**Figure 9.**
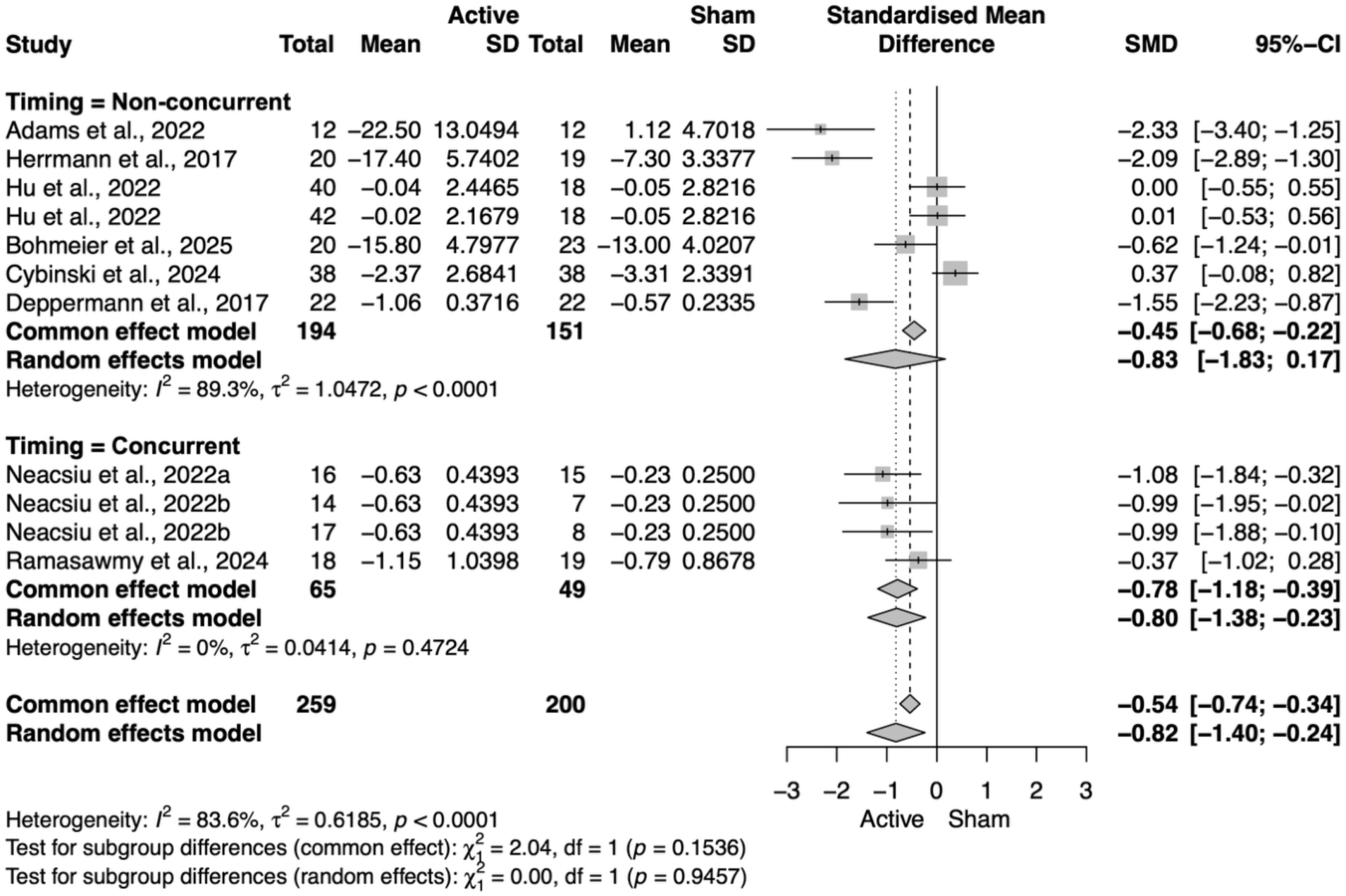
Forest plot for subcategory of NIBS timing on anxiety outcomes.

## 4 Discussion

This meta-analysis provides a comprehensive quantitative synthesis of randomized controlled trials examining the efficacy of NIBS combined with evidence-based psychotherapy for psychiatric disorders. Our findings demonstrate that active NIBS paired with psychotherapy produces significantly greater improvement in primary clinical symptoms compared to sham NIBS with psychotherapy, with a small-to-moderate overall effect size (SMD = -0.38). Critically, our moderator analyses reveal that the magnitude and significance of this benefit depend on specific implementation parameters: NIBS modality, temporal alignment of stimulation with therapy, diagnostic category, and the psychotherapy delivery method. These findings offer mechanistic insights into how brain state modulation enhances therapeutic learning and provide a framework for optimizing combined treatment protocols.

### 4.1 Primary findings and clinical significance

The overall effect size we observed (SMD = -0.38) aligns with prior meta-analytic estimates reporting small-to-moderate benefits of combined approaches (He et al., 2022; Xu et al., 2023). However, our results diverge from these earlier syntheses in critical ways. First, by restricting inclusion to trials that compared active NIBS plus psychotherapy against sham NIBS plus psychotherapy—thereby controlling for nonspecific effects of psychotherapy—we isolate the specific augmentation effect attributable to brain stimulation. Second, by requiring clinician-verified evidence-based psychotherapy, we minimize heterogeneity introduced by non-manualized or poorly validated interventions. This stringent choice reduces confounding but also narrows the evidence base, underscoring the limited number of well-controlled trials in this emerging field.

Our subgroup analyses demonstrate that rTMS, but not tDCS, yielded statistically significant improvement when combined with psychotherapy. This divergence may reflect mechanistic differences between modalities. TMS directly induces action potentials and modulates cortical excitability with high spatial precision, whereas tDCS shifts membrane potentials subthreshold, producing weaker and more diffuse neuromodulatory effects. The greater efficacy of rTMS could also be attributed to more established clinical protocols, higher dose consistency across trials, and more robust sham control procedures. However, the tDCS subgroup included fewer studies and exhibited substantial heterogeneity, which may have precluded detection of smaller effects. Emerging home-based tDCS protocols, recently FDA-approved for major depressive disorder (Bikson et al., 2026), may offer unique advantages for scalability and adherence that warrant further investigation in combination paradigms.

We found no significant effects of combined NIBS and psychotherapy on executive functioning (n = 9 studies, SMD = 0.12) or quality of life (n = 9 studies, SMD = 0.16). This null finding should be interpreted cautiously given the limited sample and heterogeneity in assessment instruments. Prior meta-analyses have reported inconsistent results: Fu et al. (2025) found no cognitive benefits of rTMS in depression across multiple domains (Fu et al., 2025), while Xu et al. (2023) observed improvement in global cognition but not quality of life (Xu et al., 2023). Future trials should continue assessing these outcomes to clarify whether combined treatment impacts cognitive function and wellbeing.

### 4.2 The critical role of temporal alignment

Perhaps the most theoretically and clinically important finding from this meta-analysis is the moderating effect of NIBS timing. Non-concurrent delivery, where stimulation preceded or followed psychotherapy, produced significant symptom reduction (SMD = -0.56), whereas concurrent delivery did not (SMD = -0.11). This pattern held specifically for rTMS delivered non-concurrently (SMD = -0.47), with no significant effects for concurrent rTMS, concurrent tDCS, or non-concurrent tDCS. These findings appear to contradict prevailing theories that emphasize state-dependent plasticity, which predict maximal benefit when stimulation co-occurs with active cognitive or emotional engagement (Luber and Lisanby, 2014; Sathappan et al., 2019).

Several mechanisms may account for the superiority of non-concurrent protocols. First, priming paradigms—in which stimulation precedes therapy—may enhance synaptic plasticity and metaplasticity, rendering treatment-engaged circuits more responsive to subsequent therapeutic learning (Tatti et al., 2022). Preclinical work demonstrates that TMS can upregulate brain-derived neurotrophic factor (BDNF) and long-term potentiation, creating a neurobiological window of heightened plasticity that extends beyond the stimulation session itself (Gersner et al., 2011). Second, consolidation paradigms—in which stimulation follows therapy—may stabilize newly formed memory traces through reconsolidation mechanisms, strengthening extinction learning or cognitive restructuring achieved during psychotherapy (Saccenti et al., 2024). Third, concurrent delivery may introduce practical barriers that undermine psychotherapy fidelity. The percussive noise, somatic sensations, and attentional demands of concurrent TMS can interfere with mindfulness practice, exposure tolerance, or therapeutic alliance, as evidenced by high dropout rates in some concurrent protocols (Cavallero et al., 2021; Demina et al., 2025).

Importantly, our secondary analysis of anxiety outcomes revealed a different pattern: concurrent NIBS significantly reduced anxiety symptoms (SMD = -0.80, 95% CI [-1.38; -0.23]), while non-concurrent administration showed a comparable point estimate (SMD = -0.83) but with wide confidence intervals that did not reach statistical significance (95% CI [-1.83; 0.17]). This may reflect limited power rather than a true mechanistic difference, as the effect sizes were nearly identical. However, the significant concurrent finding suggests that anxiety reduction may particularly benefit from real-time neuromodulation during exposure or emotional processing, when anxious arousal is actively generated. In contrast, broader clinical improvement—captured by general outcome measures—showed the opposite pattern, with significant effects only for non-concurrent delivery (SMD = -0.56). This dissociation suggests that optimal timing may be symptom-specific: anxiety scales may capture state-dependent reactivity that responds to concurrent modulation (Spielberger, 1983), whereas primary outcomes assess stable symptom severity and functional impairment that may require neuroplastic consolidation beyond the therapy session. Future trials should systematically compare timing schedules within the same protocol with adequate power to disentangle these effects.

### 4.3 Psychotherapy factors: Modality, delivery, and treatment integrity

Among psychotherapy modalities, only CBT combined with NIBS produced statistically significant improvement (SMD = -0.48), while exposure therapy and mindfulness did not. This finding likely reflects both the larger sample of CBT trials and the heterogeneity within the exposure- and mindfulness-based therapies. Exposure-based interventions, which constituted nearly half of all studies, included protocols ranging from brief symptom provocation (as in the FDA-cleared OCD protocol) to full multi-session exposure and response prevention. Similarly, mindfulness interventions varied from brief audio-guided exercises to structured mindfulness-based stress reduction programs. The CBT category, by contrast, encompassed manualized protocols with clearer structural consistency across trials.

Our analysis also revealed that only human-delivered psychotherapy—as opposed to computerized or hybrid formats—resulted in significant augmentation by NIBS. This finding underscores the importance of the therapeutic relationship, real-time adaptation to patient responses, and the clinician’s ability to navigate complex emotional states that may arise during treatment. Computerized interventions, while offering standardization and scalability, may lack the interpersonal depth necessary to capitalize on NIBS-induced plasticity. These programs often present abbreviated or simplified versions of evidence-based techniques, which may not fully engage the cognitive and emotional processes targeted by NIBS. Hybrid approaches, which combine human oversight with digital content delivery, showed intermediate but non-significant effects, suggesting a potential middle ground worthy of further refinement.

Treatment integrity, encompassing therapist adherence to manualized protocols and competent delivery, is a critical but under-reported dimension in this literature. Our clinician screening identified that only 39.3% of studies employed fully evidence-based, manualized psychotherapy, with the remainder using only partial elements of validated treatments. Moreover, only three studies (10.7%) explicitly reported adherence monitoring or competence ratings. This gap is concerning because treatment integrity is consistently associated with superior clinical outcomes in psychotherapy research (Esposito et al., 2024; Power et al., 2022). When psychotherapy is diluted or poorly implemented, the synergistic potential of combined NIBS-psychotherapy protocols may be undermined. Future trials must prioritize treatment integrity by using manualized interventions, training therapists to fidelity, and systematically monitoring adherence and competence throughout the study.

### 4.4 Diagnostic considerations and transdiagnostic potential

At the diagnostic level, significant improvement was observed only for anxiety disorders (SMD = -0.70), with no significant effects for addiction, OCD, pain, or PTSD. This pattern may reflect disorder-specific neural mechanisms, sample size limitations, or protocol optimization. Anxiety disorders—particularly social anxiety, specific phobias, and panic disorder—are characterized by hyperactivation of threat-detection circuits involving the amygdala, insula, and medial prefrontal cortex (Etkin and Wager, 2007), which are amenable to prefrontal neuromodulation during exposure-based interventions. The integration of NIBS with exposure therapy may reduce anticipatory anxiety, enhance extinction learning (Raij et al., 2018), or modulate dlPFC-mediated anxious arousal (Balderston et al., 2020), mechanisms that align well with the neurobiological targets of prefrontal TMS.

The absence of significant effects in PTSD is particularly notable given that six studies included PTSD populations, comparable to the anxiety subgroup. PTSD involves complex disruptions in fear extinction, emotion regulation, and memory reconsolidation (Liberzon and Abelson, 2016; Pitman et al., 2012), processes that theoretically should benefit from NIBS-augmented psychotherapy. However, PTSD trials in our sample exhibited high heterogeneity in trauma types, stimulation parameters, and psychotherapy formats, which may have obscured benefits. Additionally, PTSD is often comorbid with depression, dissociation, and chronic hyperarousal (Brady et al., 2000; Flory and Yehuda, 2015), which may require more intensive or prolonged combined treatment (Schnurr et al., 2024) than the protocols tested to date.

The limited representation of MDD in our sample (only two studies) is striking, given that MDD is the most extensively studied indication for NIBS monotherapy (Lefaucheur et al., 2020). Most combination trials in MDD have compared active NIBS plus psychotherapy against active NIBS alone, rather than against sham NIBS plus psychotherapy, and were therefore excluded by our eligibility criteria. This design choice reflects a pragmatic clinical question (does adding psychotherapy improve outcomes over stimulation alone?) but precludes conclusions about whether NIBS augments psychotherapy specifically. Future research should address this gap by testing whether TMS enhances established psychotherapies like behavioral activation or interpersonal therapy in MDD, particularly for patients with partial response to either modality alone.

### 4.5 Limitations

Several limitations warrant consideration when interpreting these findings. First, the relatively small number of eligible trials (n = 28) limited statistical power for subgroup and interaction analyses, particularly for underrepresented diagnoses such as MDD, BPD, and transdiagnostic samples. Confidence intervals were wide for several moderators, and null findings may reflect insufficient power rather than true absence of effects. Second, substantial heterogeneity (high *I*²) indicates variability across trials in stimulation parameters (frequency, intensity, target, number of sessions), psychotherapy protocols (dose, fidelity, delivery format), and patient characteristics (symptom severity, treatment resistance, medication status). While we conducted moderator analyses to parse this heterogeneity, unmeasured confounders likely remain. Third, we faced constraints in categorizing NIBS timing. We originally intended to distinguish between stimulation delivered before, during, or after psychotherapy to enable more specific mechanistic recommendations. However, many studies did not specify exact timing beyond reporting non-concurrent delivery, resulting in insufficient sample sizes for separate before/after categories. Consequently, we collapsed all non-concurrent protocols into a single category. While this approach maintained statistical power, it precluded differentiation between priming effects (pre-session stimulation) and consolidation effects (post-session stimulation), which may operate through distinct neuroplastic mechanisms. Future trials should explicitly report whether NIBS precedes or follows therapy to enable more granular temporal analyses.

Fourth, treatment integrity was under-reported. Only 10.7% of studies documented therapist adherence or competence, making it difficult to determine whether psychotherapy was delivered as intended. The majority of studies (60.7%) used only partial elements of evidence-based protocols rather than fully manualized treatments, introducing additional heterogeneity. Without systematic fidelity monitoring, it is impossible to disentangle whether null findings reflect ineffective combinations or poor psychotherapy implementation. Fifth, the majority of studies allowed concurrent psychotropic medications, which may interact with NIBS effects. Benzodiazepines, for instance, can blunt TMS-induced plasticity through GABAergic mechanisms, while antipsychotics may alter cortical excitability. Although medication status was reported in most studies, few conducted sensitivity analyses to evaluate its moderating role. Finally, our analysis focused on short-term outcomes, typically assessed immediately post-treatment or within a few weeks. Evidence for the durability of combined NIBS-psychotherapy effects is limited. Long-term follow-up data—at 6, 12, or 24 months—were reported in only a subset of trials, and we lacked sufficient data to conduct meta-analytic synthesis of maintenance effects. This is a critical gap, as one of the theoretical advantages of combining psychotherapy with NIBS is the potential to enhance durability through consolidation of therapeutic learning. Future trials should prioritize extended follow-up periods to determine whether augmentation by NIBS produces more sustained remission than psychotherapy alone.

### 4.6 Future directions

Future research should also prioritize mechanistic endpoints. Incorporating neuroimaging (resting-state and task-based fMRI), electrophysiology (EEG, TMS-evoked potentials), and peripheral biomarkers (BDNF, inflammatory cytokines) can illuminate how combined treatment alters neural circuits and whether neuroplastic changes mediate clinical outcomes. Biomarker-stratified trials could identify which patients are most likely to benefit from combined treatment, moving the field toward precision psychiatry. fMRI-individualized TMS targeting may also allow for more precise manipulation of diagnosis-specific neural circuits of interest, a method that was only used in approximately 11% of included studies. Additionally, researchers should explore novel NIBS modalities not yet tested in combination with psychotherapy, including tACS, tFUS, and closed-loop neurostimulation paradigms that adapt in real time to brain state.

Finally, implementation science research is critical to translate these findings into real-world practice. Combined NIBS-psychotherapy protocols are resource-intensive, requiring specialized equipment, trained operators, and extended treatment schedules. Cost-effectiveness analyses, scalability studies, and dissemination trials are needed to determine whether combined approaches can be feasibly integrated into routine clinical care, particularly in underserved settings. The recent FDA approval of home-based tDCS for depression raises intriguing possibilities for remotely supervised combined treatment, pairing home-delivered stimulation with teletherapy. Such models could dramatically expand access while maintaining treatment fidelity.

## 5 Conclusion

This meta-analysis demonstrates that noninvasive brain stimulation, when combined with evidence-based psychotherapy, produces significant improvement in psychiatric symptoms beyond the effects of psychotherapy with sham stimulation. However, this benefit is conditional on critical implementation parameters. rTMS delivered non-concurrently with psychotherapy— either before or after sessions—emerges as the most effective strategy, particularly when paired with human-delivered, manualized CBT for anxiety disorders. The superiority of non-concurrent delivery challenges prevailing state-dependent models and suggests that priming or consolidation mechanisms may be more important than real-time co-activation. Treatment integrity, including the use of fully evidence-based psychotherapy and fidelity monitoring, is essential but under-reported in the current literature.

While these findings provide an evidence-based framework for optimizing combined treatment protocols, significant gaps remain. The field needs larger, mechanistically informed trials with long-term follow-up, standardized reporting of psychotherapy fidelity, individualized NIBS targeting, and biomarker-guided patient selection. By addressing these challenges, the integration of brain stimulation with psychotherapy holds promise as a next-generation approach to enhance the speed, magnitude, and durability of therapeutic change in psychiatric care.

## Data Availability

All data produced in the present work are contained in the manuscript and the supplementary information, as well as in the original study manuscripts analyzed in this meta-analysis.

## Acknowledgements

Zhi-De Deng, Lysianne Beynel, Ethan Greenstein, Eudora Jones, Neil Baker, Sunday M. Francis, Bruce Luber, and Sarah H. Lisanby are supported by the National Institute of Mental Health (NIMH) Intramural Research Program (ZIAMH002955). The contributions of the NIH authors are considered Works of the United States Government. The findings and conclusions presented in this paper are those of the authors and do not necessarily reflect the views of the NIH or the U.S. Department of Health and Human Services.

