## Supplementary Information for "Noninvasive brain stimulation combined with evidence-based psychotherapy for psychiatric disorders: A meta-analysis of optimal implementation parameters"

### Supplementary Material

This supplementary material provides additional detail on the meta-analysis of noninvasive brain stimulation (NIBS) combined with evidence-based psychotherapy for psychiatric disorders. The contents include:

- **Supplementary Figures S1–S5:** Forest plots stratified by key moderator variables, including diagnostic category, psychotherapy type, level of evidence supporting psychotherapy, administration method, and interactions between NIBS modality and psychotherapy category.
- **Supplementary Table S1:** Detailed study population characteristics and outcome measures for all 28 included randomized controlled trials (31 independent comparison groups).
- **Supplementary Table S2:** NIBS parameters including modality (rTMS or tDCS), timing relative to psychotherapy, anatomical target, targeting method, stimulation intensity, protocol specifications, and number of sessions.
- **Supplementary Table S3:** Psychotherapy parameters including therapy type (CBT, exposure, mindfulness), delivery method (human, computerized, or hybrid), concurrent medication allowance, adherence monitoring, number of sessions, and session duration.

All analyses used random-effects models to account for between-study heterogeneity. Standardized mean differences (SMD) are reported with 95% confidence intervals, with negative values indicating superiority of active NIBS combined with psychotherapy over sham NIBS combined with psychotherapy.

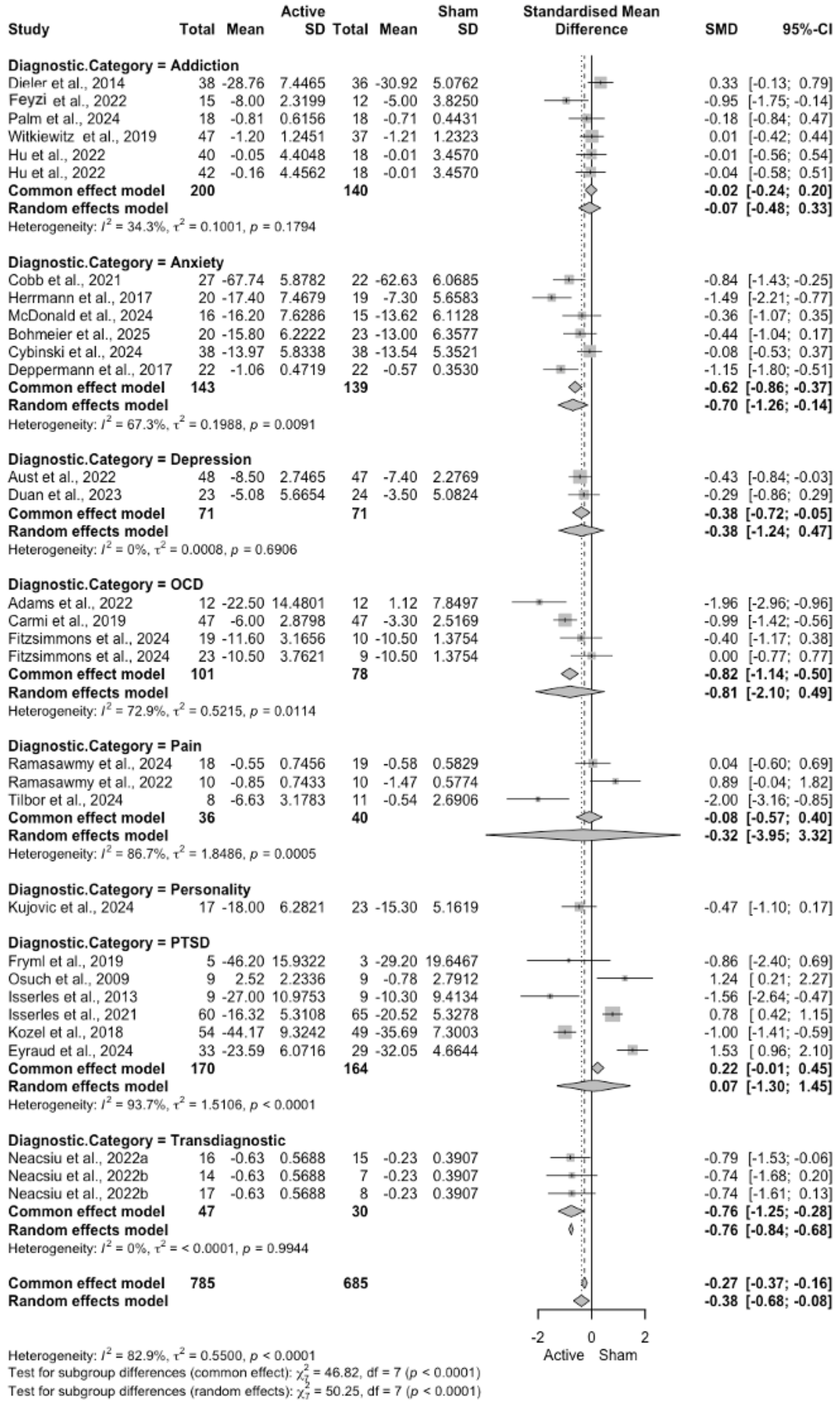


Figure S1: **Primary outcome forest plot stratified by diagnostic category.** Standardized mean differences (SMD; negative values favor active NIBS + psychotherapy) are shown for each trial with 95% confidence intervals, pooled within diagnostic categories using random-effects models. Significant effects were observed only for anxiety disorders ($\mathrm{SMD} = -0.70$, 95% CI: [-1.26; $-0.14$], $p < 0.05$). No significant effects were found for addiction, depression, OCD, pain, personality disorders, PTSD, or transdiagnostic samples. Heterogeneity ($I^2$) and $p$-values are reported for each diagnostic subgroup. The overall pooled effect across all diagnoses ($\mathrm{SMD} = -0.38$, 95% CI: $[-0.68; -0.08]$) is shown at the bottom.

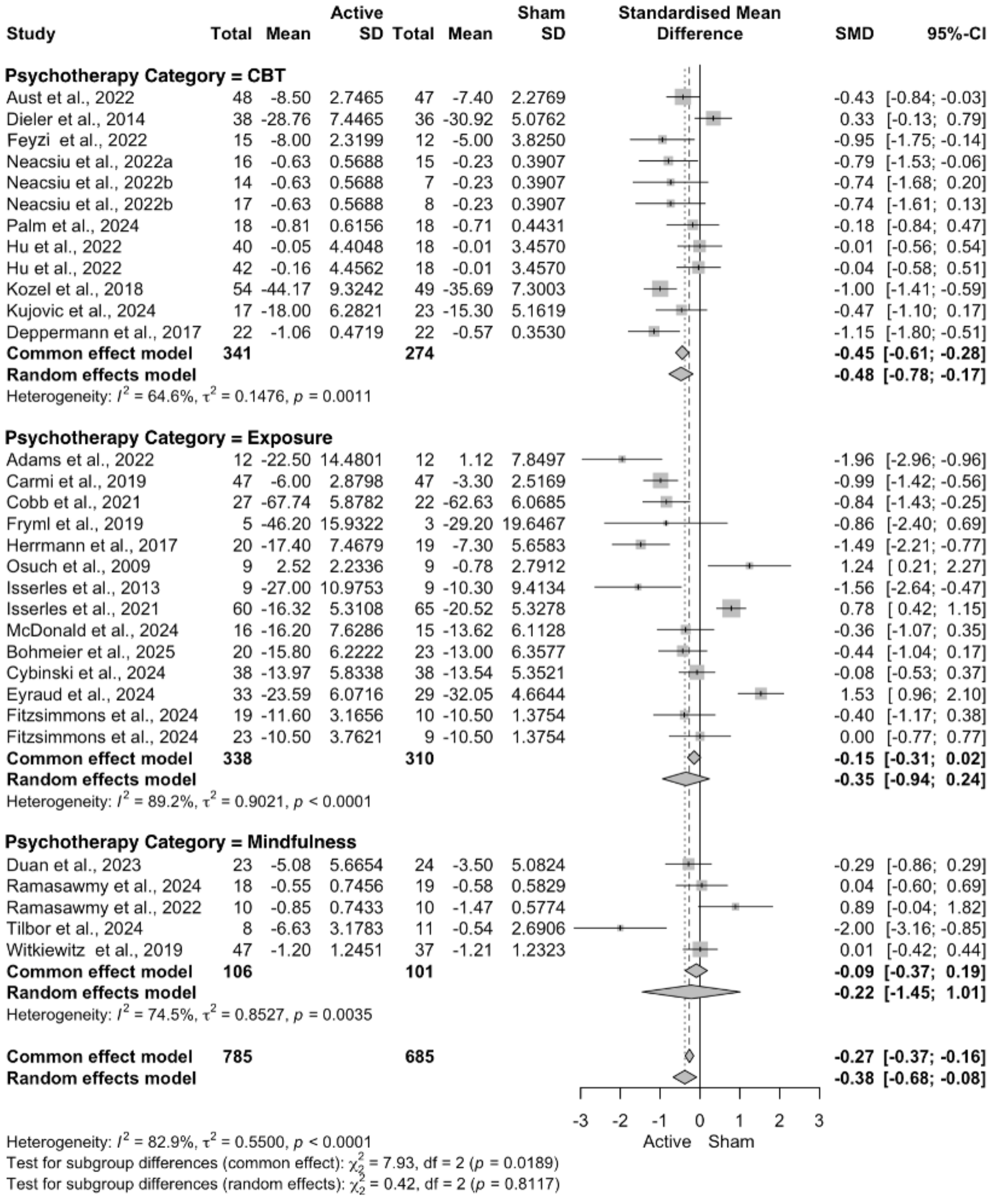


Figure S2: **Primary outcome forest plot stratified by psychotherapy category.** SMDs pooled within psychotherapy categories. Cognitive behavioral therapy (CBT) showed significant improvement (SMD $==-0.48$, 95% CI: $[-0.78; -0.17]$), while exposure therapy (SMD $=-0.35$, 95% CI: $[-0.94; 0.24]$) and mindfulness (SMD $=-0.22$, 95% CI: $[-1.45; 1.01]$) did not.

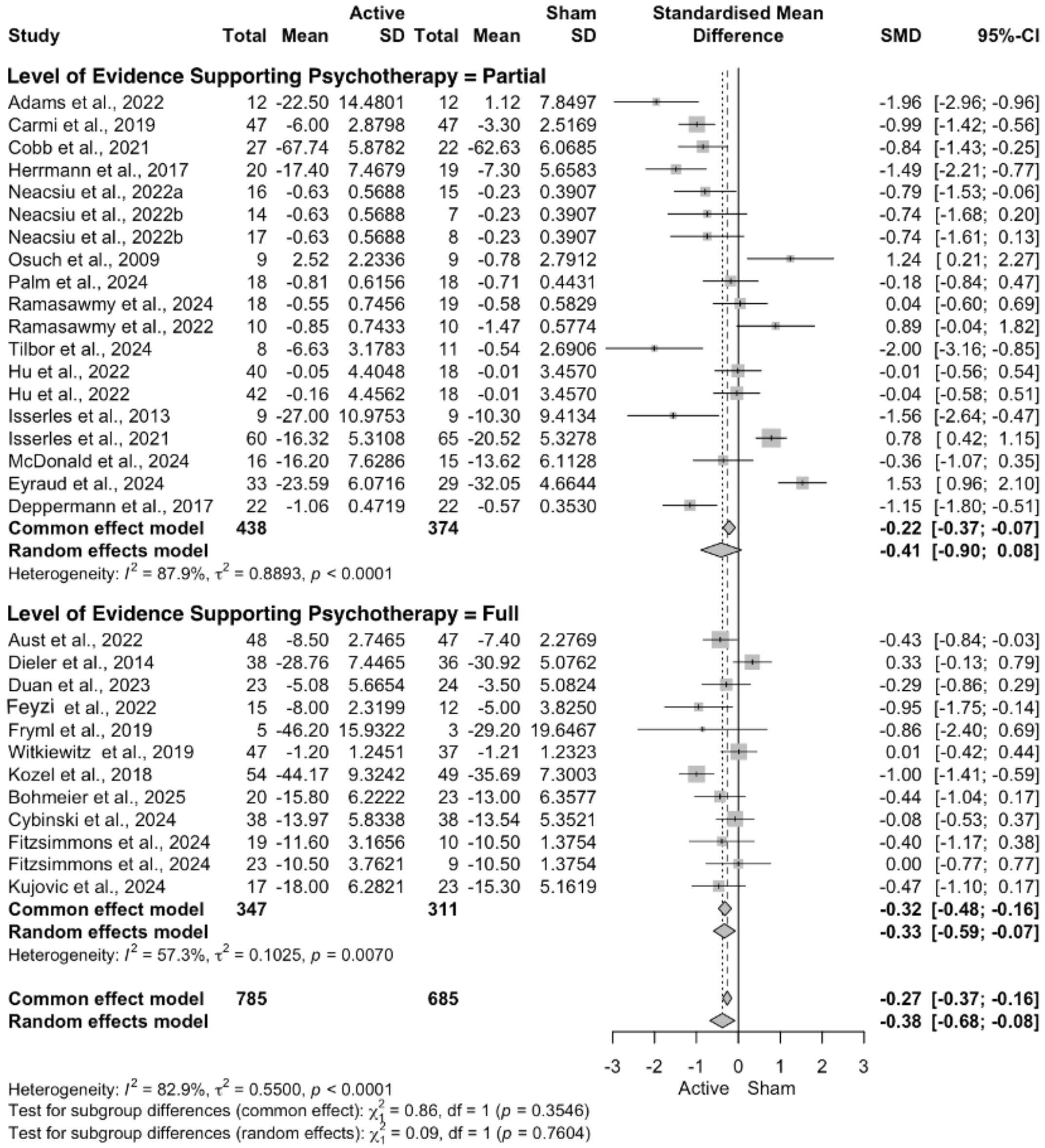


Figure S3: **Primary outcome forest plot stratified by level of evidence supporting the psychotherapy.** SMDs are pooled by whether studies used fully evidence-based psychotherapy (manualized protocols with empirical support) or only partial elements of evidence-based approaches. Although both fully evidence-based ($\mathrm{SMD} = -0.33$, 95% CI: $[-0.59;\ -0.07]$) and partial evidence-based ($\mathrm{SMD} = -0.41$, 95% CI: $[-0.90;\ 0.08]$) psychotherapy showed trends toward improvement, only the fully evidence-based group reached statistical significance.

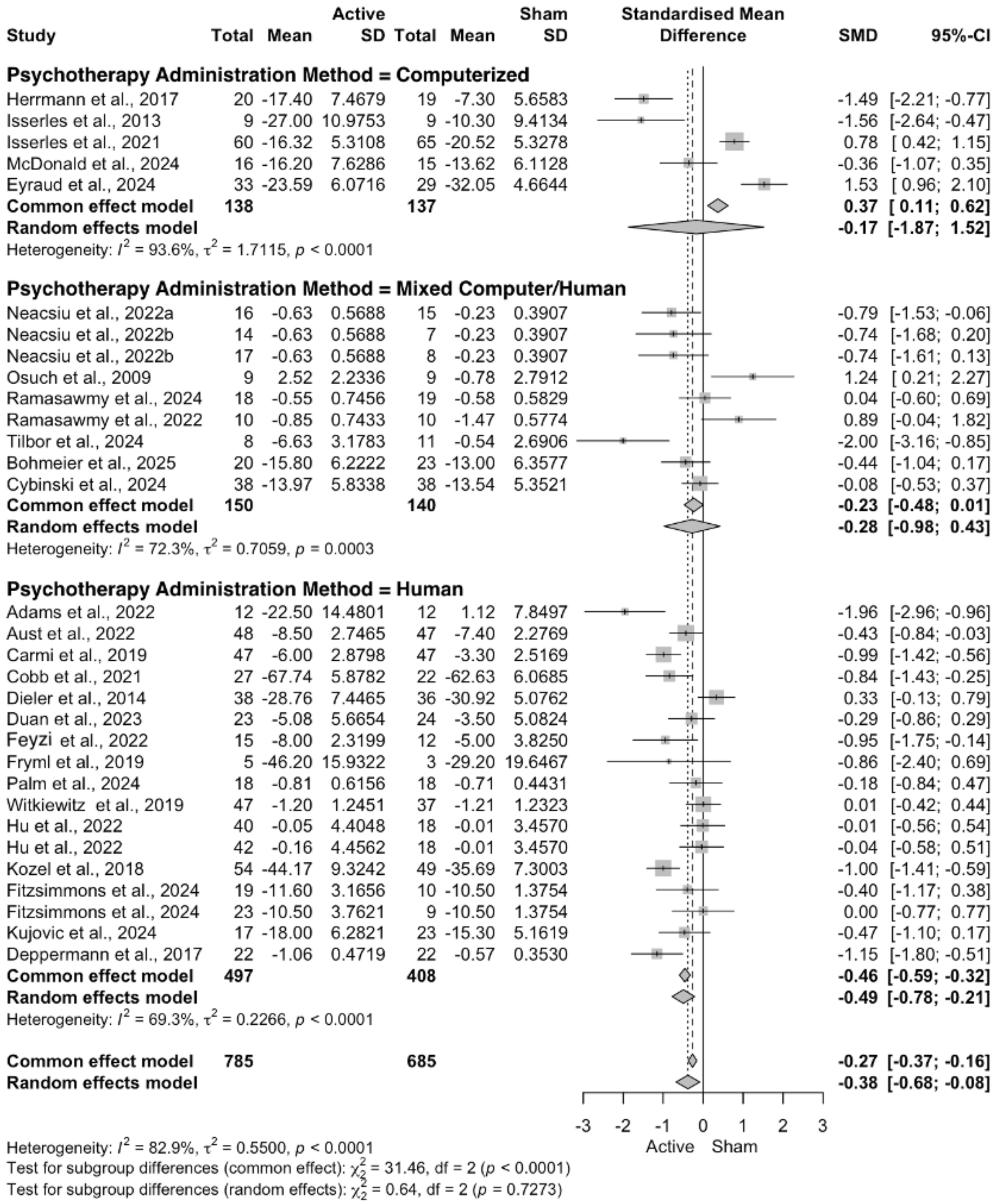


Figure S4: **Primary outcome forest plot stratified by psychotherapy administration method.** SMDs pooled by delivery format. Only human-administered psychotherapy showed significant improvement (SMD $= -0.49$, 95% CI: $[-0.78; -0.21]$), while computerized delivery (SMD $= -0.17$, 95% CI: $[-1.87; 1.52]$) and mixed computer/human formats (SMD $= -0.28$, 95% CI: $[-0.98; 0.43]$) did not reach statistical significance.

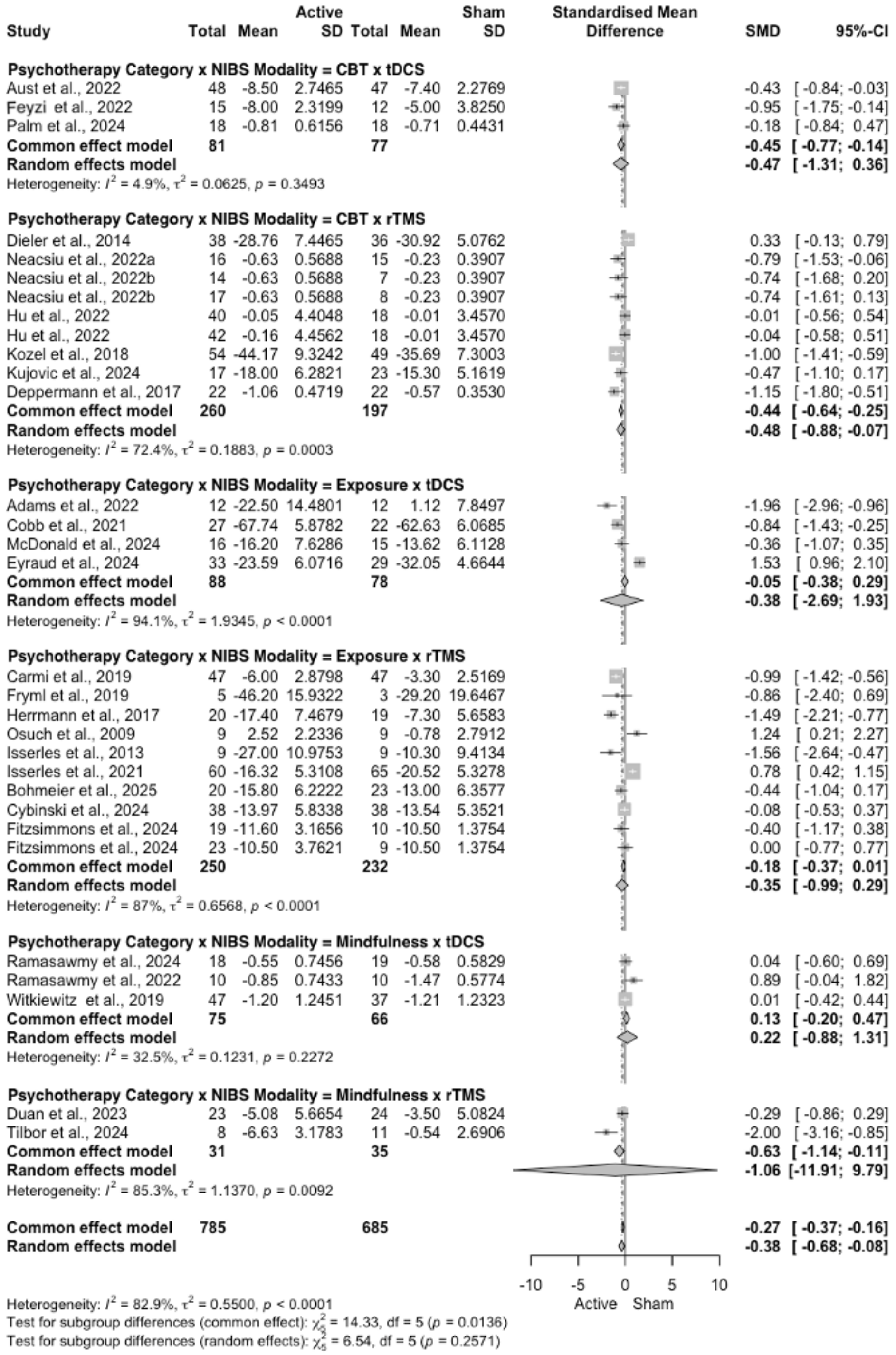


Figure S5: **Primary outcome forest plot for the interaction between NIBS modality and psychotherapy category.** SMDs pooled by the combination of NIBS modality (rTMS or tDCS) and psychotherapy type (CBT, exposure, or mindfulness). Among the combinations with sufficient data for analysis, only CBT paired with rTMS showed significant improvement ($\mathrm{SMD} = -0.48$, 95% CI: $[-0.88; -0.07]$). No significant effects were observed for CBT + tDCS ($\mathrm{SMD} = -0.47$, 95% CI: $[-1.31; 0.36]$), exposure + tDCS ($\mathrm{SMD} = -0.38$, 95% CI: $[-2.69; 1.93]$), exposure + rTMS ($\mathrm{SMD} = -0.35$, 95% CI: $[-0.99; 0.29]$), mindfulness + tDCS ($\mathrm{SMD} = 0.22$, 95% CI: $[-0.88; 1.31]$), or mindfulness + rTMS ($\mathrm{SMD} = -1.06$, 95% CI: $[-11.91; 9.79]$).

Table S1: Study population and outcomes

| **Study** | ***N*** **(active / sham)** | **Mean age (y) (active / sham)** | **Diagnosis** | **Primary outcome** | **Secondary: Depression** | **Secondary: Anxiety** | **Secondary: QoL** | **Secondary: Executive function** |
|---|---|---|---|---|---|---|---|---|
| Adams et al. 2022 [1] | 12 / 12 | 32.4 / 34.4 | OCD | SUDS | | | | |
| Aust et al. 2022 [2] | 43 / 42 | 42.7 / 39.0 | MDD | MADRS | | | SF-36-MH | |
| Bohmeier et al. 2025 [3] | 20 / 23 | 40.9 / 49.9 | Anxiety | AQ (ACRO subscale) | ADS-K | AQ (avoidance) | | |
| Carmi et al. 2019 [4] | 47 / 47 | 41.1 / 36.5 | OCD | YBOCS | HAM-D | | SDS | |
| Cobb et al. 2021 [5] | 27 / 22 | 21.6 / 20.7 | Anxiety | BAT 1 | | | | |
| Cybinski et al. 2024 [6] | 38 / 38 | 37.8 / 41.6 | Anxiety | AQ (ACRO subscale) | ADS-K | ASI-3 | | |
| Deppermann et al. 2017 [7] | 22 / 22 | 37.6 / 36.3 | Anxiety | PAS–Observer Rated | | | | |
| Dieler et al. 2014 [8] | 38 / 36 | 46.7 / 46.3 | Addiction | SER (Temptation) | | | | |
| Duan et al. 2023 [9] | 23 / 24 | 58.3 / 53.6 | MDD | HAM-D | | | MBI | MMSE |
| Eyraud et al. 2024 [10] | 30 / 27 | 31.5 / 35.6 | PTSD | CAPS-5 | BDI | STAI-A | WHOQoL-BREF | Stroop, N-back |
| Fayaz Feyzi et al. 2022 [11] | 15 / 12 | 34.0 / 37.0 | Addiction | OCDUS | | | | WCST |
| Fitzsimmons et al. [12] Group 1 | 23 / 19 | 31.5 / 40.3 | OCD | YBOCS | BDI | | | |
| Fitzsimmons et al. [12] Group 2 | 19 / 19 | 40.3 / 40.3 | OCD | YBOCS | BDI | | | |
| Fryml et al. 2019 [13] | 5 / 3 | 27.0 / 30.0 | PTSD | CAPS | HAM-D | HAM-A | | |
| Herrmann et al. 2017 [14] | 20 / 19 | 43.2 / 46.6 | Anxiety | AQ (anxiety) | ADS | | | |
| Hu et al. 2022 [15] Group 1 | 42 / 37 | 44.0 / 46.0 | Addiction | OCDS | PHQ-9 | GAD-7 | PSQI | MoCA |
| Hu et al. 2022 [15] Group 2 | 40 / 37 | 48.0 / 46.0 | Addiction | OCDS | PHQ-9 | GAD-7 | PSQI | MoCA |
| Isserles et al. 2013 [16] | 9 / 9 | 49.0 / 40.4 | PTSD | CAPS | HAM-D | | | |
| Isserles et al. 2021 [17] | 40 / 51 | 44.8 / 43.7 | PTSD | CAPS-5 | | | | |
| Kozel et al. 2018 [18] | 32 / 30 | 34.1 / 32.9 | PTSD | CAPS | QIDS | | | IPF |
| Kujovic et al. 2024 [19] | 17 / 23 | 24.8 / 25.5 | BPD | BSL-23 | BDI | | | GAF |
| McDonald et al. 2024 [20] | 16 / 15 | 19.8 / 20.0 | Anxiety | PRPSA | | | | |
| Neacsiu et al. 2022a [21] | 16 / 15 | 32.9 / 34.7 | Transdiagnostic | DERS | | ERQ | OQ-45 | WSAS |
| Neacsiu et al. 2022b [22] Group 1 | 14 / 15 | 33.3 / 29.5 | Transdiagnostic | DERS | | ERQ | OQ-45 | WSAS |
| Neacsiu et al. 2022b [22] Group 2 | 17 / 15 | 27.8 / 29.5 | Transdiagnostic | DERS | | ERQ | OQ-45 | WSAS |
| Osuch et al. 2009 [23] | 9 / 9 | 41.4 / 41.4 | PTSD | CAPS | HAM-D | | | |
| Palm et al. 2024 [24] | 18 / 18 | 51.4 / 50.6 | Addiction | QSU Factor 1 | | | | |
| Ramasawmy et al. 2022 [25] | 10 / 10 | 59.7 / 48.2 | Pain | NRS | DASS-21 | DASS-21 | FIQ | |
| Ramasawmy et al. 2024 [26] | 18 / 19 | 51.8 / 51.6 | Pain | NRS | DASS-21 | DASS-21 | FIQ | |
| Tilbor et al. 2024 [27] | 8 / 11 | 51.7 / 46.2 | Pain | SF-MPQ | HAM-D | | | |
| Witkiewitz et al. 2019 [28] | 47 / 37 | 51.4 / 53.4 | Addiction | Drinks per day | | | | SST |

ADS: Allegemeine Depressionsskala (English: eneral Depression Scale); ADS-K: Kurzform der Allegemeine Depressionsskala (English: Short form of the General Depression Scale); AQ: Acrophobia Questionnaire; ASI: Anxiety and Sensitivity Index; BAT-1/BAT-2: Behavioral Avoidance Test; BDI: Beck Depression Inventory; BPD: Borderline personality disorder; BSL-23: Borderline Symptom List; CAPS-5: Clinician Administered PTSD Scale; DASS: Depression Anxiety Stress Scales; DERS: Difficulties in Emotion Regulation Scale; EGF/GAF: Global Assessment of Functioning Scale (French/English); ERQ: Emotion Regulation Questionnaire; FIQ: Fibromyalgia Impact Questionnaire; GAD-7: Generalized Anxiety Disorder 7-Item; HAM-A: Hamilton Anxiety Rating Scale; HAM-D: Hamilton Depression Rating Scale; IPF: Inventory of Psychosocial Functioning; MADRS: The Montgomery–Åsberg Depression Rating Scale; MBI: Modified Barthel Index; MDD: Major depressive disorder; MMSE: Mini-Mental State Examination; MoCA: Montreal Cognitive Assessment; NRS: Numerical Rating Scale; OCD: Obsessive-compulsive disorder; OCDS: Obsessive Compulsive Drinking Scale; OQ-45: Outcome Questionnaire; PAS–Observer Rated: Panic and Agoraphobia Scale; PHQ-9: Patient Health Questionnaire; PQSI: Pittsburgh Sleep Quality Index; PRPSA: The Personal Report of Public Speaking Anxiety; PTSD: Post-traumatic stress disorder; QIDS: Quick Inventory of Depressive Symptomology; QSU Factor 1: Questionnaire on Smoking Urges (Factor 1 – intention and desire to smoke); SDS: Sheehan Disability Scale; SER: Questionnaire of Self-Efficacy in Smokers; SF-36-MH: Short-Form 36 – Mental Health; SF-MPQ: Short-form McGill Pain Questionnaire; SST: Stop Signal Task; STAI-A: State-Trait Anxiety Inventory for Adults; SUDS: The Subjective Units of Distress Scale; WCST: Wisconsin Card Sorting Test; WMS: Weschler Memory Scale; WHOQOL-BREFL: World Health Organization Quality of Life Brief; WSAS: The Work and Social Adjustment Scale; Y-BOCS: Yale-Brown Obsessive Compulsive Scale

### Table S2: NIBS parameters

| Study | NIBS | NIBS-therapy timing | Target | Targeting method | Intensity | TMS protocol | No. NIBS sessions | NIBS duration (min) |
|---|---|---|---|---|---|---|---|---|
| Adams et al. 2022 [1] | tDCS | Non-concurrent | Anode over medial PFC | Scalp/EEG | 1.5 mA | – | 1 | 20 |
| Aust et al. 2022 [2] | tDCS | Concurrent | Anode over left PFC | Scalp/EEG | 2 mA | – | 12 | 30 |
| Cobb et al. 2021 [5] | tDCS | Non-concurrent | Anode over medial PFC | Scalp/EEG | 1.7 mA | – | 1 | 20 |
| Eyraud et al. 2024 [10] | tDCS | Concurrent | Anode over left PFC | Scalp/EEG | 2 mA | – | 10 | 20 |
| Fayaz Feyzi et al. 2022 [11] | tDCS | Non-concurrent | Anode over left PFC | Scalp/EEG | 2 mA | – | 16 | 20 |
| McDonald et al. 2024 [20] | tDCS | Concurrent | Anode over medial PFC | Scalp/EEG | 2 mA | – | 1 | 20 |
| Palm et al. 2024 [24] | tDCS | Non-concurrent | Anode over left PFC | Scalp/EEG | 2 mA | – | 5 | 20 |
| Ramasawmy et al. 2022 [25] | tDCS | Concurrent | Anode over left M1 | Scalp/EEG | 2 mA | – | 10 | 20 |
| Ramasawmy et al. 2024 [26] | tDCS | Concurrent | Anode over left M1 | Scalp/EEG | 2 mA | – | 10 | 20 |
| Witkiewitz et al. 2019 [28] | tDCS | Concurrent | Anode over right PFC | Scalp/EEG | 2 mA | – | 8 | 30 |
| Bohmeier et al. 2025 [3] | TMS | Non-concurrent | Left PFC | Scalp/EEG | 80% RMT | iTBS | 2 | 3 |
| Carmi et al. 2019 [4] | TMS | Concurrent | Medial PFC | Scalp/EEG | 100% RMT | 20 Hz | 29 | 18 |
| Cybinski et al. 2024 [6] | TMS | Non-concurrent | Left PFC | sMRI | 100% RMT | iTBS | 2 | 3 |
| Deppermann et al. 2017 [7] | TMS | Non-concurrent | Left PFC | Scalp/EEG | 80% RMT | iTBS | 15 | 3 |
| Dieler et al. 2014 [8] | TMS | Non-concurrent | Right PFC | Scalp/EEG | 80% RMT | iTBS | 4 | 3 |
| Duan et al. 2023 [9] | TMS | Concurrent | Left PFC | Scalp/EEG | 80% RMT | 10 Hz | 20 | 20 |
| Fitzsimmons et al. 2025 [12] Group 1 | TMS | Non-concurrent | Left PFC | fMRI | 110% RMT | 10 Hz | 16 | 20 |
| Fitzsimmons et al. 2025 [12] Group 2 | TMS | Non-concurrent | Left M1 | fMRI | 110% RMT | 10 Hz | 16 | 20 |
| Fryml et al. 2019 [13] | TMS | Concurrent | Left or right PFC | Scalp/EEG | 120% RMT | 10 Hz | 5 | 30 |
| Herrmann et al. 2017 [14] | TMS | Non-concurrent | Medial PFC | Scalp/EEG | 100% RMT | 10 Hz | 2 | 20 |
| Hu et al. 2022 [15] Group 1 | TMS | Non-concurrent | Right PFC | Scalp/EEG | 110% RMT | 10 Hz | 10 | 12.5 |
| Hu et al. 2022 [15] Group 2 | TMS | Non-concurrent | Left PFC | Scalp/EEG | 110% RMT | 10 Hz | 10 | 12.5 |
| Isserles et al. 2013 [16] | TMS | Non-concurrent | Medial PFC | Scalp/EEG | 120% RMT | 20 Hz | 12 | 15.5 |
| Isserles et al. 2021 [17] | TMS | Non-concurrent | Medial PFC | Scalp/EEG | 100% RMT | 18 Hz | 12 | 30 |
| Kozel et al. 2018 [18] | TMS | Non-concurrent | Right PFC | Scalp/EEG | 110% RMT | 1 Hz | 12 | 30 |
| Kujovic et al. 2024 [19] | TMS | Non-concurrent | Left PFC | Scalp/EEG | 80% RMT | iTBS | 20 | 3 |
| Neacsiu et al. 2022a [21] | TMS | Concurrent | Left PFC | fMRI | 120% RMT | 10 Hz | 1 | 40 |
| Neacsiu et al. 2022b [22] Group 1 | TMS | Concurrent | Left PFC | Scalp/EEG | 120% RMT | 10 Hz | 1 | 40 |
| Neacsiu et al. 2022b [22] Group 2 | TMS | Concurrent | Right PFC | Scalp/EEG | 120% RMT | 10 Hz | 1 | 40 |
| Osuch et al. 2009 [23] | TMS | Concurrent | Right PFC | Scalp/EEG | 100% RMT | 1 Hz | 20 | 30 |
| Tilbor et al. 2024 [27] | TMS | Non-concurrent | Medial PFC | Scalp/EEG | 100% RMT | 20 Hz | 20 | 18 |

sMRI: structural MRI; fMRI: functional MRI; M1: primary motor cortex; RMT: resting motor threshold; PFC: prefrontal cortex

Table S3: Therapy parameters

| Study | Therapy | Delivery | Concurrent medication allowed | Adherence completed | No. therapy sessions | Therapy duration per session (min) |
|---|---|---|---|---|---|---|
| Aust et al. 2022 [2] | CBT | Human | Yes | Yes | 12 | 100 |
| Deppermann et al. 2017 [7] | CBT | Human | Yes | No | 11 | 90 |
| Dieler et al. 2014 [8] | CBT | Human | No | No | 6 | 90 |
| Fayaz Feyzi et al. 2022 [11] | CBT | Human | No | No | 24 | 60 |
| Hu et al. 2022 [15] | CBT | Human | No | No | 8 | 60 |
| Kozel et al. 2018 [18] | CBT | Human | Yes | No | 12 | 60 |
| Kujovic et al. 2024 [19] | CBT | Human | Yes | Yes | 8 wks total (1–2 sessions/wk) | Variable |
| Neacsiu et al. 2022a [21] | CBT | Hybrid | Yes | No | 1 | 36 |
| Neacsiu et al. 2022b [22] | CBT | Hybrid | Yes | No | 1 | 36 |
| Palm et al. 2024 [24] | CBT | Human | No | No | 5 | 5 |
| Adams et al. 2022 [1] | Exposure | Human | Yes | No | 2 | 50 |
| Bohmeier et al. 2025 [3] | Exposure | Hybrid | No | No | 2 | 27.3 |
| Carmi et al. 2019 [4] | Exposure | Human | Yes | No | 29 | 3–5 |
| Cobb et al. 2021 [5] | Exposure | Human | Yes | Yes | 1 | 30 |
| Cybinski et al. 2024 [6] | Exposure | Hybrid | No | No | 2 | 28.9 |
| Eyraud et al. 2024 [10] | Exposure | Computerized | Yes | No | 10 | 20 |
| Fitzsimmons et al. 2025 [12] | Exposure | Human | Yes | No | 20 | 60 |
| Fryml et al. 2019 [13] | Exposure | Human | Yes | Unclear | 5 | 40 |
| Herrmann et al. 2017 [14] | Exposure | Computerized | No | Unclear | 2 | 50 |
| Isserles et al. 2013 [16] | Exposure | Computerized | Yes | No | 12 | 4 |
| Isserles et al. 2021 [17] | Exposure | Computerized | Yes | No | 12 | 1.5 |
| McDonald et al. 2024 [20] | Exposure | Computerized | Yes | No | 1 | 18 |
| Osuch et al. 2009 [23] | Exposure | Hybrid | Yes | No | 20 | 5 |
| Duan et al. 2023 [9] | Mindfulness | Human | Yes | No | 6 | 60 |
| Ramasawmy et al. 2022 [25] | Mindfulness | Hybrid | Yes | No | 10 | 30 |
| Ramasawmy et al. 2024 [26] | Mindfulness | Hybrid | Yes | No | 10 | 20 |
| Tilbor et al. 2024 [27] | Mindfulness | Hybrid | Yes | No | 20 | 20 |
| Witkiewitz et al. 2019 [28] | Mindfulness | Human | No | No | 8 | 120 |

CBT: cognitive behavioral therapy

[21] A. D. Neacsiu, L. Beynel, J. L. Graner, S. T. Szabo, L. G. Appelbaum, M. J. Smoski, and K. S. LaBar. Enhancing

cognitive restructuring with concurrent fMRI-guided neurostimulation for emotional dysregulation - A randomized controlled trial. *J Affect Disord*, 301:378–389, 2022. doi: 10.1016/j.jad.2022.01.053.
